# Increasing concentration of COVID-19 by socioeconomic determinants and geography in Toronto, Canada: an observational study

**DOI:** 10.1101/2021.04.01.21254585

**Authors:** Sharmistha Mishra, Huiting Ma, Gary Moloney, Kristy CY Yiu, Dariya Darvin, David Landsman, Jeffrey C. Kwong, Andrew Calzavara, Sharon Straus, Adrienne K Chan, Effie Gournis, Heather Rilkoff, Yiqing Xia, Alan Katz, Tyler Williamson, Kamil Malikov, Rafal Kustra, Mathieu Maheu-Giroux, Beate Sander, Stefan D Baral, on behalf of the COVID-19 Heterogeneity Research Group

**Affiliations:** St. Michael’s Hospital, Unity Health Toronto, Toronto, Canada; Division of Infectious Diseases, Department of Medicine, University of Toronto, Toronto, Canada; ICES, Toronto, Canada; Public Health Ontario, Toronto, Canada; Department of Family and Community Medicine, Faculty of Medicine, University of Toronto, Toronto, Canada; Dalla Lana School of Public Health, University of Toronto, Toronto, Canada; Department of Medicine, St. Michael’s Hospital, University of Toronto, Toronto, Canada; Division of Infectious Diseases, Sunnybrook Health Sciences, University of Toronto, Toronto, Canada; Institute of Health Policy, Management and Evaluation, University of Toronto, Toronto, Canada; Toronto Public Health, City of Toronto, Toronto, Canada; Department of Epidemiology, Biostatistics and Occupational Health, School of Population and Global Health, McGill University, Montréal, Canada; Departments of Community Health Sciences and Family Medicine, Rady Faculty of Health Sciences, University of Manitoba, Winnipeg, Canada; Department of Community Health Sciences, University of Calgary, Calgary, Canada; Centre for Health Informatics, University of Calgary, Calgary, Canada; Capacity Planning and Analytics Division, Ontario Ministry of Health, Toronto, Canada; Department of Epidemiology, Johns Hopkins School of Public Health, Baltimore, United States

## Abstract

**Background:** Inequities in the burden of COVID-19 observed across Canada suggest heterogeneity within community transmission.

**Objectives:** To quantify the magnitude of heterogeneity in the wider community (outside of long-term care homes) in Toronto, Canada and assess how the magnitude in concentration evolved over time (January 21 to November 21, 2020).

**Design:** Retrospective, population-based observational study using surveillance data from Ontario’s Case and Contact Management system.

**Setting:** Toronto, Canada.

**Participants:** Laboratory-confirmed cases of COVID-19 (N=33,992).

**Measurements:** We generated epidemic curves by SDOH and crude Lorenz curves by neighbourhoods to visualize inequities in the distribution of COVID-19 cases by social determinants of health (SDOH) and estimated the crude Gini coefficient. We examined the correlation between SDOH using Pearson correlation coefficients.

**Results:** The Gini coefficient of cumulative cases by population size was 0.41 (95% CI: 0.36-0.47) and were estimated for: household income (0.20, 95%CI: 0.14-0.28); visible minority (0.21, 95%CI: 0.16-0.28); recent immigration (0.12, 95%CI: 0.09-0.16); suitable housing (0.21, 95%CI: 0.14-0.30); multi-generational households (0.19, 95%CI: 0.15-0.23); and essential workers (0.28, 95% CI: 0.23-0.34). Most SDOH were highly correlated.

Locally acquired cases were concentrated in higher income neighbourhoods in the early phase of the epidemic, and then concentrated in lower income neighbourhoods. Mirroring the trajectory of epidemic curves by income, the Lorenz curve shifted over time from below to above the line of equality with a similar pattern across SDOH.

**Limitations:** Study relied on area-based measures of the SDOH and individual case counts of COVID-19. We cannot infer concentration of cases by specific occupational exposures given limitation to broad occupational categories.

**Conclusion:** COVID-19 is increasingly concentrated by SDOH given socioeconomic inequities and structural racism.

**Primary Funding Source:** Canadian Institutes of Health Research.

## INTRODUCTION

Inequities in the burden of COVID-19 were observed early in Canada(1) and around the world,(2-4) suggesting that racialized and economically marginalized communities faced disproportionate risks of acquisition.

Descriptive data suggested a temporal shift in how SARS-COV-2 may have spread through transmission networks over a short period of time within cities.(5, 6) For example, in the Greater Toronto Area, Canada, (population over 7 million)(7) the first wave from January to August, 2020 quickly divided into micro-epidemics in congregate settings (e.g., long-term care homes) and what seemed to be more diffuse spread in the wider community.(6) However, an examination of cumulative cases suggested considerable heterogeneity within the wider community with greater risks associated with household size and occupation.(8)

Heterogeneity is an established marker of inequities, with the application of Lorenz curves and Gini coefficients increasingly used to quantify and compare health inequities(9) in infectious diseases.(10-12) Health inequities, in turn, represent unmet prevention needs acting as mechanistic drivers of onward transmission in epidemics.(13, 14) Specifically, observed cases concentrated within a smaller subset of the population which often indicates disproportionate risks of onward transmission, whether at the individual-level mediated via higher contact rates(13, 14) or more commonly, at the network-level such as household density or high-exposure occupations.(8, 15)

Gini coefficients have been commonly used in sexually transmitted infections as a measure of spatial concentration to help identify spatial ‘core-groups’ that experience disproportionate risks in the context of core-group theory.(10, 11) Core-group theory is based on the premise that if prevention efforts are not effective within a smaller group of people experiencing very high risks of both acquisition and onward transmission, the epidemic cannot be controlled (10, 11). In addition to sexually transmitted infections, Lorenz curves and Gini coefficients have been used for a range of other health outcomes.(16-19) For example, inequities in longevity over time,(18) oral health,(17) and premature mortality(20) have been examined against income-levels, and more recently inequities in health outcomes have been examined across other social determinants, including unemployment.(16)

The present study’s overarching objective is to improve our understanding of the role of social determinants in shaping SARS-CoV-2 transmission dynamics in Toronto, the largest city and economic capital of Canada. Specifically, we first quantified the magnitude of heterogeneity in the wider community (outside of long-term care homes) by comparing the concentration of cases by area-level social determinants. Second, we assessed how the magnitude in concentration of COVID-19 cases evolved from January to November, 2020.

## METHODS

### Study design, setting, and population

We conducted a retrospective, population-based observational study using routinely collected surveillance data; reported according to the STROBE (reporting of observational studies) guidelines.(21) The study population comprised the City of Toronto (population 2,731,571).(22) The study period included cases with a January 21 (first documented case was reported on January 23, 2020)(23) to November 21, 2020 episode date. The episode date is either the date of symptom-onset as self-reported by individuals, or the test-submission date in cases where symptom data are either missing or individuals were asymptomatic.(24) We excluded individuals residing in long-term care homes because congregate settings represent a different epidemic setting from the wider community. Our unit of analysis was the census geographic unit of the dissemination area (DA), which represents on average 400-700 residents.(25)

### Data sources

We used Ontario’s Case and Contact Management (CCM+) surveillance system of person-level, anonymized, surveillance data which includes information on laboratory-confirmed cases by episode date, reported date, the DA of residence, and demographic, exposure, and setting-specific characteristics (e.g., -long-term care residence). The DA of residence was determined using the *Postal Code Conversion File* version-SLI.(26)

For DA-level variables on the social determinants, we used publicly available data from the 2016 Canadian Census,(27) and two variables that were not publicly available: after-tax, per-person equivalent income ranking across DAs within the Toronto which was obtained from ICES (a not-for-profit research institute that securely houses Ontario’s health-related data); and a measure of multi-generational households which was curated by and sourced from the Ontario Community Health Profiles Partnership.(28)

### Measures

For social determinants, we considered measures previously shown to be associated with test-positivity;(29) and mechanistically drive transmission dynamics by considering frequency of contacts and who contacts whom.(30) These include:

1. *socio-demographic* indicators that are proxies of economic barriers and systemic racism (income; % visible minority; % recent immigration);
2. *dwelling-related* variables (% suitable housing;(31, 32) % multi-generational households); and
3. *occupation-related* variables (% working in essential services [health; trades, transport and equipment operation; sales and services; manufacturing and utilities; resources, agriculture, and production])(33)

Details and definitions for each variable are included in **Appendix Table A1**.

### Analyses

We generated crude Lorenz curves by DA to visualize inequalities in the distribution of confirmed SARS-CoV-2 cases. Lorenz curves are used to assess the relationship between two cumulative distributions: the cumulative proportion of diagnoses and the cumulative proportion of the population. This information can be summarized using the estimated crude Gini coefficient (the comparison of the cumulative proportion of population against the cumulative proportion of diagnoses) where a coefficient of zero represents complete equality and one represent complete inequality. We included all DAs in this analysis, including DAs without SARS-CoV-2 cases. We excluded cases with missing DA numbers that we were not able to link to DA level characteristics.

We examined the correlation between social determinants using Pearson correlation coefficients.(34) To investigate concentration of confirmed cases by social determinants, we first described the cumulative and daily epidemic curves by each social determinant. We then generated Lorenz curves using x-axes that represent the proportion of the population ranked by DA-level estimates of each value of the social determinant and subsequently estimated the crude Gini coefficient, over the entire study period and for each mutually exclusive time-period using the dates of large-scale restriction measures in Toronto: January to March 16 (pre-closure/shutdown); March 17 to May 18 (during shutdown and prior to stage 1 re-opening); May 19 to June 23 (stage 1); June 24 to July 30 (stage 2); July 31 to October 9 (stage 3); October 10 to November 21 (modified stage-2) (**Appendix Table A2**).(35) We generated 95% confidence intervals for the Lorenz curves and Gini coefficients using bootstrapping (9, 11).

All analyses were conducted in R (version 4.0.2), and spatial maps were generated using ArcGIS (version 10.7).

### Ethics approval

The *University of Toronto Health Sciences Research Ethics Board* (protocol no. 39253) approved the study.

## RESULTS

Between January 21 and November 21, 2020, there were N=33,992 observed cases of COVID-19, excluding long-term care residents (**Appendix Figure A1**). The median population size of a DA was 450 (interquartile range, 446-768, **Appendix Table A3**). Among 3,702 DAs in the City of Toronto, 404 DAs had zero reported cases, representing 10.9% of DAs and 8.1% of the population. Data on social determinants were missing in 0.51-0.57% of DAs (**Table 1**). The proportion of cases that were travel-acquired decreased over time: from 51% (pre-closure/shutdown) to 3% (during shutdown), 1% (stage 1), 5% (stage 2), 3% (stage 3), and 1% (modified stage-2).

**Table 1.** Gini coefficients of cumulative COVID-19 cases*, by area-level measures in Toronto, Canada from January 21, 2020 to November 21, 2020.

|  |  |  | Cases (N=33,992)* |  |
| --- | --- | --- | --- | --- |
|  | % of dissemination areas not included because missing information on the social determinant | Median (IQR) across all DAs | Gini coefficient | 95% CI |
| Population size | N/A | 450 (446 - 768) | 0.41 | 0.36-0.47 |
| **Socio-demographic** |  |  |  |  |
| Household income | 0 | 45749.5 (37815.8 - 56389.2) | 0.20 | 0.14-0.28 |
| % visible minority | 0.51 | 42.0 (22.8 – 69.0) | 0.21 | 0.16-0.28 |
| % recent immigration | 0.51 | 3.9 (1.9 - 7.7) | 0.12 | 0.09-0.16 |
| **Dwelling-related** |  |  |  |  |
| % suitable housing | 0.51 | 92.6 (85.9 - 96.6) | 0.21 | 0.14-0.30 |
| % multi-generational households | 0.57 | 7.6 (3.4 - 12.8) | 0.19 | 0.15-0.23 |
| **Occupation-related** |  |  |  |  |
| Essential services<sup>†</sup> | 0.51 | 43.8 (31.4 – 57.1) | 0.28 | 0.23-0.34 |

\*Excluding cases among residents of long-term care homes
<sup>†</sup> Essential services include: trades, transport, and equipment operation; sales and services; manufacturing and utilities; and resources, agriculture, and production; and healthcare.
CI: confidence interval; DA: dissemination area; IQR: interquartile range

### Geographic concentration of cases

The overall Gini coefficient of cumulative cases by population size was 0.41 (95% CI: 0.36-0.47) **(Figure 1A)**. For example, 53.7% (95% CI: 53.2-54.3%) of cumulative cases were diagnosed in 25% of the population, with the largest concentration of cases in the north-west part of the city (**Figure 1B**) which overlap with the social determinants under study (**Appendix Figure A2**).

**Figure 1.**
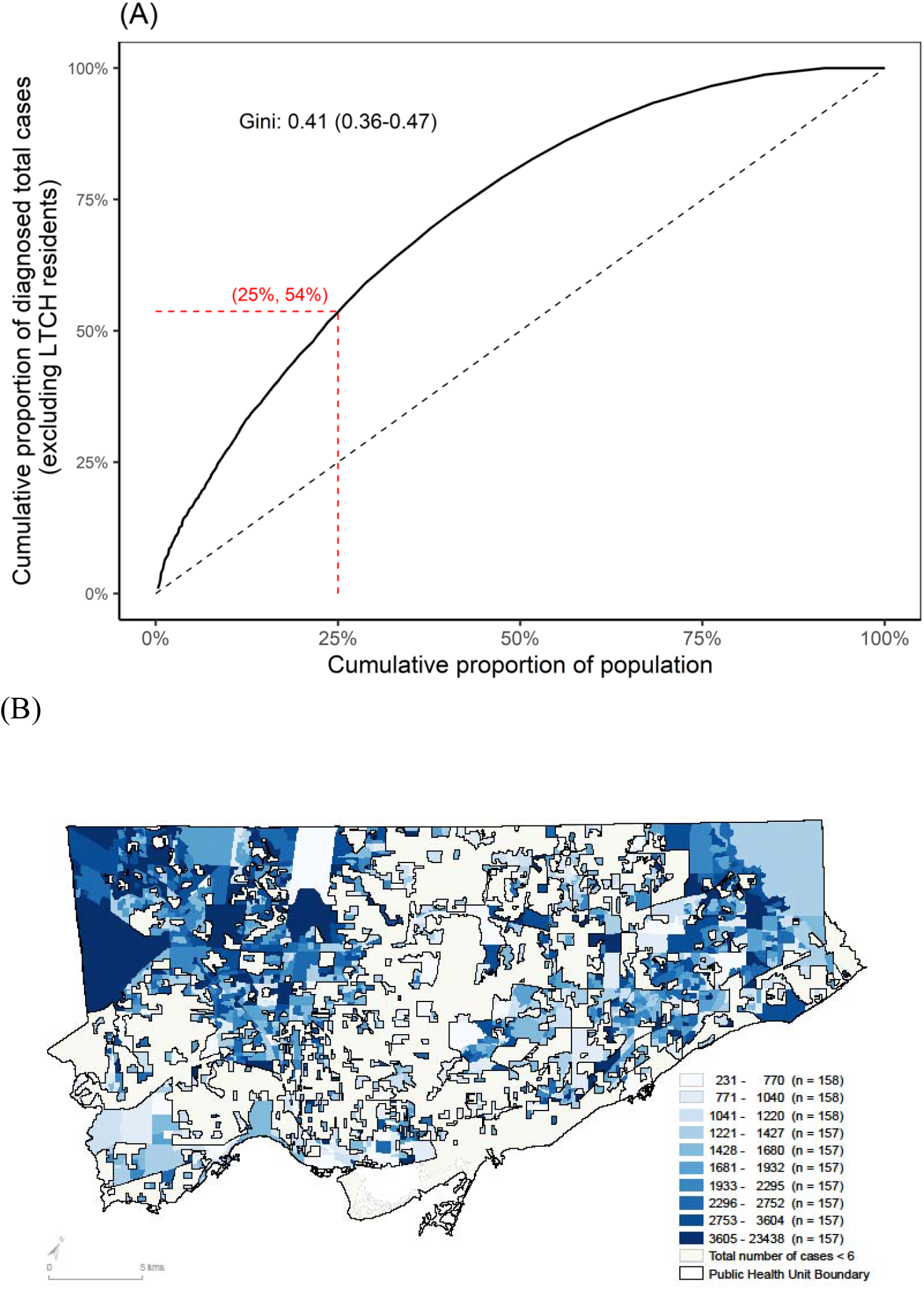
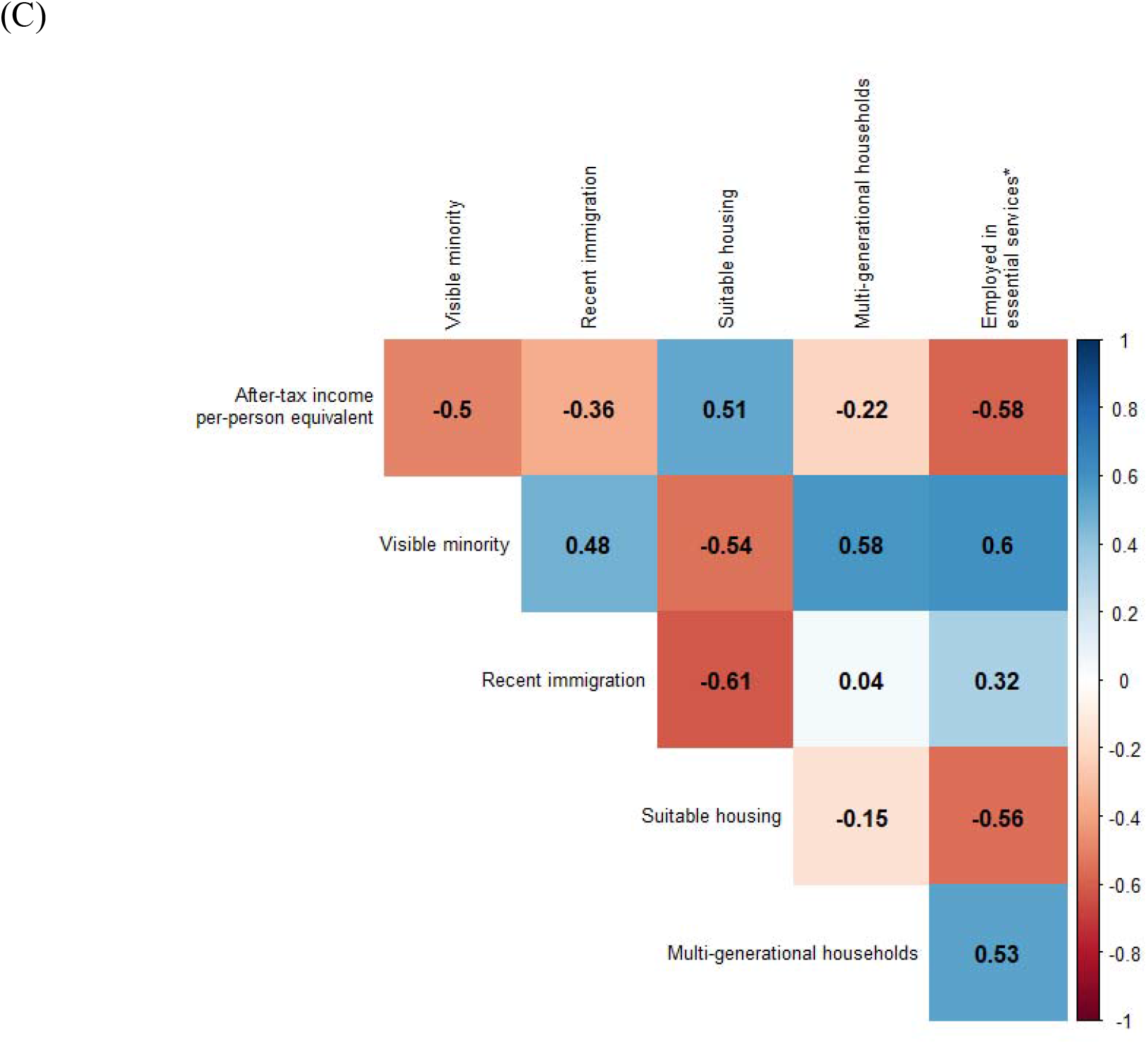
Concentration of COVID-19 cases by proportion of population in the City of Toronto, Canada (January 21 to November 21, 2020). In Panel A, the Lorenz curve depicts the cumulative proportion of laboratory-confirmed diagnoses, excluding residents of LTCH, by the cumulative proportion of the population by dissemination-area. The dashed line represents the line of equality. For example, 53.7% (95% CI: 53.2%-54.3%) of cases were diagnosed among 25% of the population as shown by the dashed horizontal and vertical red lines. Panel B shows the map of lowest to highest deciles with respect to laboratory-confirmed diagnoses per capita by dissemination-area for the City of Toronto public health unit; and excluding cases among LTCH residents. Panel C depicts a heat map of the correlation between social determinants at the dissemination area, where 1 represents a perfect positive correlation and −1 represents a perfect negative correlation. LTCH: long-term care homes

### Distribution and correlation of social determinants

At the DA-level, the median per-person equivalent after-tax income was $45,750 CAD per year; the median proportion residing in suitable housing was 92.6%; and the median proportion working in non-health essential services was 38.5% (**Table 1**). **Table A4** shows the values of each social determinant across deciles.

The correlation matrix of social determinants suggests considerable overlap (**Figure 1C)**. For example, DAs with the lowest income were correlated with a higher prevalence of essential services workers (correlation coefficient, −0.58) and, multigenerational households (correlation coefficient, −0.22), whereas higher-income DAs were correlated with a higher prevalence of suitable housing (correlation coefficient, 0.51).

### Epidemic trajectory by social determinants

The epidemic curve of cumulative cases per 100,000 population, stratified by decile for each social determinant is shown for one variable from each category (household income, suitable housing, and essential services) in **Figure 2**. Results for the remaining three social determinants are presented in **Appendix Figure A3**. The daily rates are shown as deciles in **Appendix Figure A4**.

**Figure 2.**
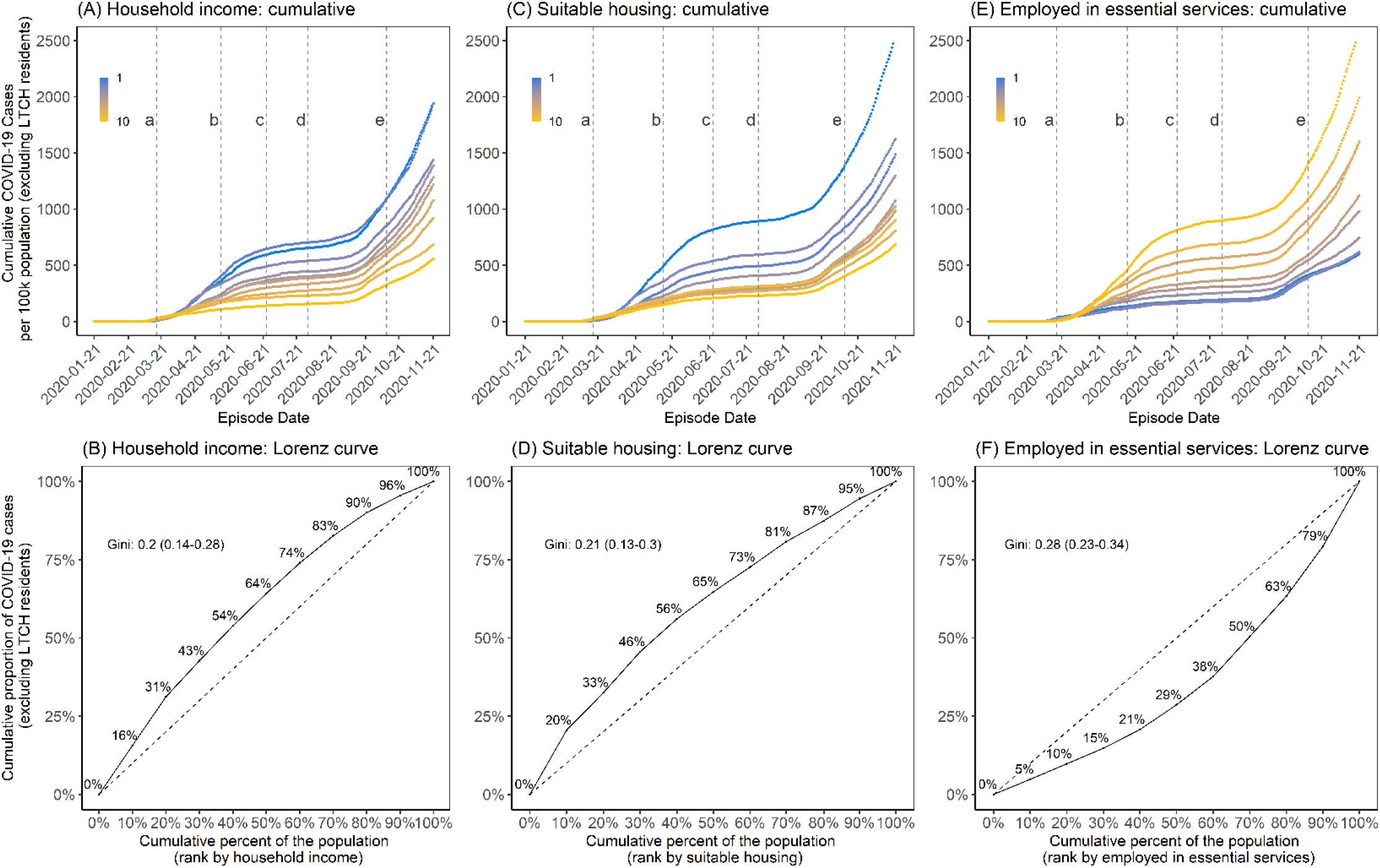
Cumulative epidemic curve and cumulative Lorenz curve by three social determinants (income, suitable housing, and essential services). Panels A and B represent income (after-tax, per-person equivalent) deciles where the lowest decile (decile 1) represents the lowest income decile and decile 10 represents the highest income decile. Panels C and D represent suitable housing deciles where the lowest decile (decile 1) represents areas with the highest proportion of homes deemed suitable housing;(27) and the highest decile (decile 10) represents areas with the lowest proportion of homes deemed suitable housing.(27) Panels E and F represent essential services where the lowest decile represents DAs with the fewest essential workers and the highest decile (decile 10) represents the highest prevalence of individuals employed in essential services. The Lorenz curves (Panels B, D and F) depict the concentration of cases by each determinant and the dashed line represents the line of equality; for example, 30% of the population residing in the lowest income areas account for 42.7% (95% CI: 42.2%-43.3%) of cumulative cases (Panel B). The time-periods are: (a) March 17, 2020, start of shutdown; (b) May 14, 2020, start of stage 1 re-opening; (c) June 24, 2020, start of stage 2 re-opening; (d) July 31, 2020, start of stage 3 re-opening; and (e) October 10, 2020, start of modified stage 2. DA: dissemination area; LTCH: long-term care homes

COVID-19 was initially concentrated in DAs represented by higher-income households before moving into lower-income DAs in early April 2020 (**Figure 3A, Appendix Figure A4A**). For the period up to May 13, 2020, travel-acquired cases were concentrated in higher-income DAs (**Figure 3B**). Locally acquired cases, however, although similarly concentrated in higher income neighbourhoods in the early phase of the epidemic, shifted concentration into lower income neighbourhoods (**Figure 3C**). A similar pattern for other social determinants was observed (**Appendix Figure A5**). In September, 2020, with the start of the second wave, there was a similar and consistent pattern of steeper epidemic curves among lower income neighbourhoods and slower growth in higher income neighbourhoods -a pattern replicated across the other social determinants (**Appendix Figures A4B-F**).

**Figure 3.**
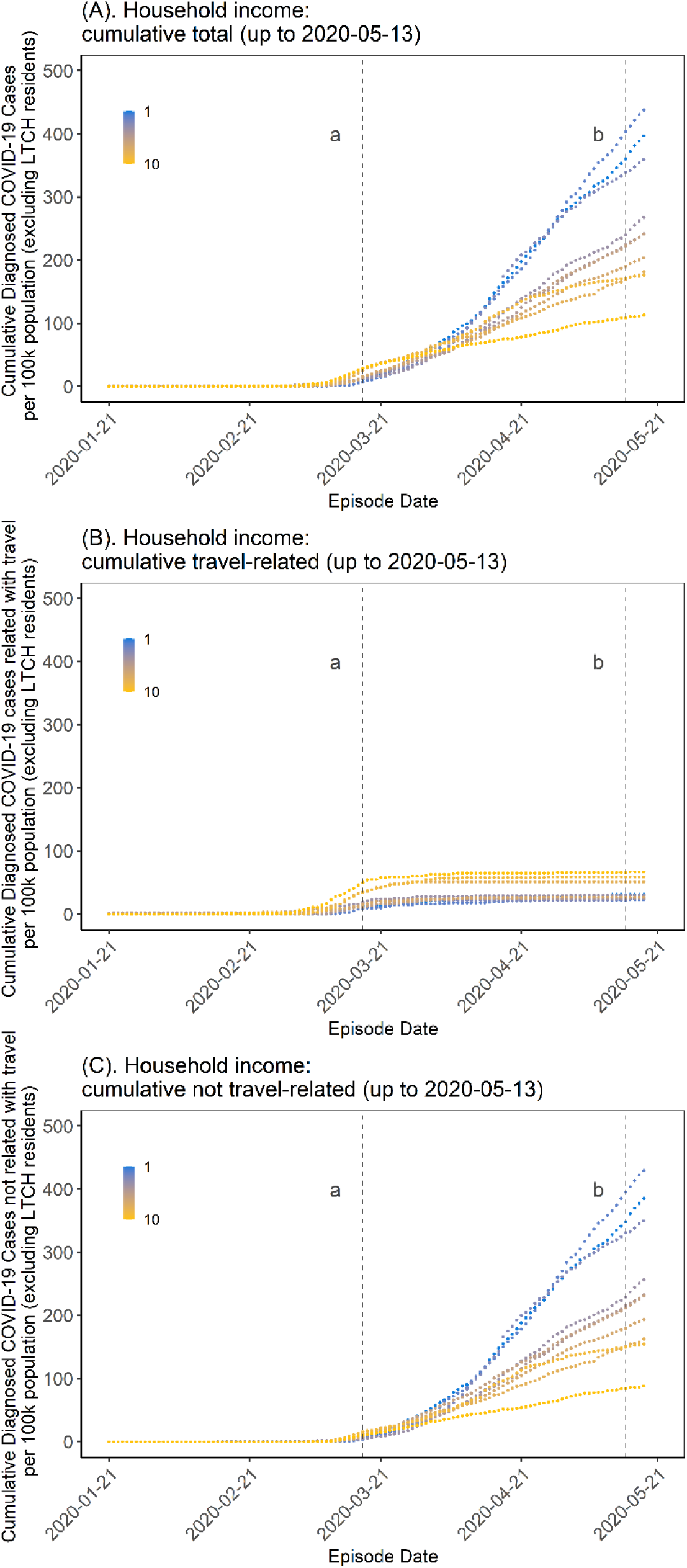
Cumulative epidemic curve by household income at the start of the COVID-19 epidemic in Toronto, Canada (January 21, 2020 to May 13, 2020). Panel A depicts all cases (excluding cases in LTCH) and demonstrates a cross-over in epidemic trajectories by DA-level income (per-person equivalent, after-tax). Early in the epidemic, per-capita rates were highest in high-income neighbourhoods but this pattern quickly reversed by early April, 2020. Panel B depicts the trajectory restricted to among travel-acquired cases which were concentrated in higher income neighbourhoods, as were the early cases before the cross-over among those not acquired through travel (Panel C). The time-periods are: (a) March 17, 2020, start of shutdown; and (b) May 14, 2020, start of stage 1 re-opening. DA: dissemination area; LTCH: long-term care homes

### Concentration of cases by social determinants

The Lorenz curves and Gini coefficients of cumulative cases for the whole study period are shown in **Figures 2B, 2D**, and **2F**; **Appendix Figures A3B, A3D**, and **A3F**, and summarized in **Table 1**. The largest Gini coefficients were estimated for essential workers (Gini coefficient 0.28, 95% CI: 0.23-0.34).

Mirroring to the trajectory of epidemic curves by income (**Figure 2A, Appendix Figure A4A**), the Lorenz curve (**Figure 4**) shifted over time from below to above the line of equality. The remainder of the Lorenz curves over time are shown in **Appendix Figures A5A-E)**, and the Gini over time is summarized in **Appendix Figure A6**. For example, the Lorenz curve for income remained below the line of equality in the early period prior the March 16 stay-at-home order (Gini 0.25, 95% CI: 0.20-0.35) when diagnosed infections were disproportionately concentrated in higher income neighbourhoods and then consistently above the line of equality thereafter when cases were disproportionately concentrated in lower income neighbourhoods.

**Figure 4.**
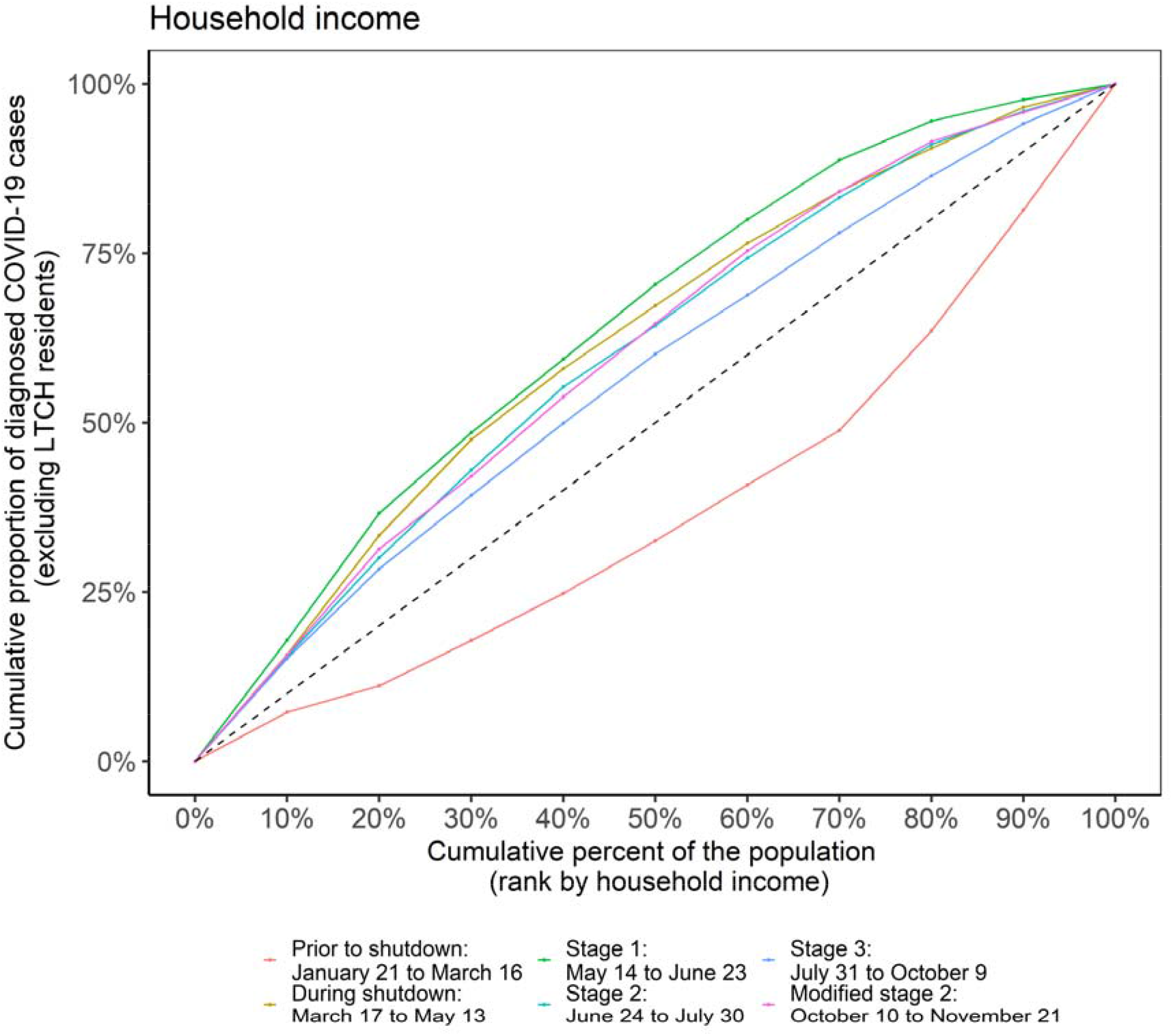
Lorenz curve and Gini coefficient of COVID-19 cases over time by income in Toronto, Canada (January 21, 2020 to November 21, 2020). The dashed line is the line of equality. The coloured solid lines represent the time periods associated with the stages of intervention: prior to shutdown (January 21, 2020 to March 16, 2020); during shutdown (March 17, 2020 to May 13, 2020); stage 1 re-opening (May 14, 2020 to June 23, 2020); stage 2 re-opening (June 24, 2020 to July 30, 2020); stage 3 re-opening (July 31, 2020 to October 9, 2020); and modified stage 2 re-opening (October 10, 2020 to November 21, 2020). Income values described are per-person equivalent and after-tax. The number of cases were initially disproportionately lower in neighbourhoods of lower income (light pink) before quickly concentrating in higher income neighbourhoods. There was less heterogeneity at the beginning of the second wave in Stage 3 (dark blue), but the epidemic concentrated again quickly by neighbourhood level income by the modified stage 2 (dark pink).

Within each time-period from the first stay-at-home order in March, the magnitude of concentration remained relatively stable during the stay-at-home orders and during Stage 1 (May-June) and Stage 2 (July-August) re-openings (**Appendix Figure A6**). However, when the second wave began in September, there was less heterogeneity across all social determinants (**Appendix Figure A6**). The Gini coefficient for income, for example, fell to 0.14 (95% CI: 0.10-0.21) before rapidly concentrating again and increasing to 0.21 (95% CI: 0.13-0.30) by the time modified restrictions resumed in October.

## DISCUSSION

Using metrics of social determinants of health, we quantified increasing inequities in COVID-19 cases over time in Toronto, Canada. We found that there has been a consistent pattern, established early, of rapid concentration of COVID-19 in small geographic areas. Areas of Toronto particularly burdened by COVID-19 have lower income, dwellings characterized by household crowding, and occupations that are not amenable to remote work.

The pattern of early travel-related cases largely concentrated in higher-income and less diverse communities with fewer essential workers are similar to that observed across the globe.(2, 15, 36-38) We also found that the early, non-travel related cases were in similar neighbourhoods (i.e., higher income) suggesting a partially assortative (“like mixes with like”) physical network with respect to income-level and other social determinants. This pattern by income suggests that early, non-travel cases initially spread within higher-income networks before entering and spreading in networks represented by lower-income households. There has been limited study of population-level mixing (or “who has contact with whom”) by income and other social determinants in the context of acute respiratory infections. For sexually transmitted infections, network assortativity has been well established within sexual networks. (39) Network assortativity for COVID-19 may be driven by the overlap between social networks and transmission networks defined by greater shared airspace and opportunities for transmission where people live and work. This finding is consistent with data on social networks suggesting variable degrees of assortativity,(40) with connections between social networks through occupation-such as caregivers.(41, 42)

The observed pattern of rapid and then persistent concentration of COVID-19 by social determinants, such as household density for example, has been consistent across high-income countries(43, 44) and in several low-and middle-income countries as well.(3, 4) Where reported, occupations not amenable to remote work have also been identified as an important risk factor for COVID-19.(15) In our study setting, essential services workers intersected with multi-generational households suggesting a connection between workplace exposures and those in home communities amplified by high rates of transmission within larger households.(45)

These results should be considered in the light of limitations. We relied on area-based measures of the social determinants of health and thus our findings may be subject to ecological fallacy. There are two reassuring elements supporting validity of these findings including the use of the smallest geographical area available (DA) and the congruence of these results with individual data characterizing race and income among people with COVID-19.(46) We were also limited to broad categories with respect to occupation, and thus cannot infer concentration of cases by specific occupational exposures. Finally, these metrics for concentration or inequities are descriptive and do not independently specify a causal mechanism.

The magnitude and direction of concentration remained relatively stable after March; that is, the Gini coefficient changed little within each intervention period. The sustained inequities are consistent with data from the United States,(47) and recently reported in Toronto,(48) which suggest that mobility-based interventions-such as wide-scale restrictions-are least likely to reduce mobility in lower-income neighbourhoods.(49, 50) Collectively, workplace mobility and the Lorenz curves by non-health essential workers suggest mobility-based interventions which restrict movement may have reached saturation of impact for a subset of the population, well before the second wave in Toronto. That is, public health measures that intend to keep people at home (e.g., lockdown or stay at home orders) may have worked on only a subset of the population and potentially reinforced disparities in COVID-19 among those who could not work remotely – especially in the absence of workplace-specific occupational health interventions.

There are several implications of the findings for public health. First, findings could be used to adapt interventions and allocate resources where transmission occurs, geographically and temporally. An example of this is vaccine allocation based on per-capita risks and not just population size: i.e. the combination of geography (hence, social determinants) and age when considering vaccination priorities to allow for the joint probability of acquisition risk and severity risk when allocating limited vaccination supply in the short-term.(51) Such efforts would also need to include active, community-tailored efforts to address vaccine confidence/trust which may be lower in communities most affected by COVID-19;(52) and account for implementation challenges with neighbourhood-specific vaccine delivery ensuring it is equitable.(53, 54) Second, the occupational and housing risks identified here may be overcome with structural interventions such as paid leave, housing supports, and alleviating barriers to testing and healthcare access. While structural interventions are often considered using a long-term horizon, immediate-term activities that are implemented at scale to those who are most at risk of acquisition and transmission are feasible and are being implemented in some settings – as evidenced by community-tailored mobile testing programs and isolation support.(55, 56) These immediate actions continue to challenge the assumption that tackling social determinants can only be addressed in the long-term while passive restrictions to reduce mobility are considered feasible by decision-makers.(57, 58) For example, immediate and actionable measures to control the spread of an infectious disease can still include active, on-the-ground support for infection prevention and control in workplaces and systematically removing barriers to sick leave to support safe, voluntary, and sustainable isolation.(57, 59) Finally, public health agencies and governments can be called upon to actively measure inequities in their COVID-19 response to better understand who their interventions are reaching and who they may be leaving behind.(60)

## CONCLUSIONS

In sum, there was a rapid epidemiologic transition in COVID-19 from an epidemic concentrated among higher income communities in Toronto related to travel to local transmission in lower income communities secondary to structural risks including structural racism often defining where people work and how they live. Similar to other infectious diseases the findings of marked inequities in COVID-19 burden in Toronto presented here suggests the existence of core-groups concentrated by social determinants of health. Specifically, these data characterized ‘core-groups’ that may be spatially-distributed, but whose underlying social determinants of health define the fundamental sources of heterogeneity in risks. Taken together, these findings suggest the added benefit of resource-based interventions to better address the specific needs of higher risk communities and thus reducing inequities in outcomes as has been advocated for by the local public health agencies.(61) Moreover, the inequities in the burden of COVID-19 appear to be sustained in the context of non-adaptive population-wide intervention strategies potentially secondary to the differential impact in reducing contact rates based on household density and occupation. Taken together, these results from Toronto suggest the potential impact of an equity-lens to inform resource-based policies and programs to optimize both the equity and effectiveness of COVID-19 intervention strategies.

## Data Availability

Reported COVID-19 cases are not publicly available and were obtained from the Public Health Ontario Integrated Public Health Information System (iPHIS) via the Ontario COVID-19 Modelling Consensus Table and with approval from the University of Toronto Health Sciences Research Ethics Board (protocol no. 39253). A subset of iPHIS data (laboratory-confirmed cases only) were made available by the Ontario Ministry of Health and Long-Term Care (MOHLTC) to the Ontario Modelling Consensus Table on a daily basis. Variables for social determinants of health are available publicly through the 2016 Canadian Census with the exception of multi-generational households and after tax income per single person equivalent (ATIPPE). Multi-generational households data is not publicly available. ATIPPE data is available through StatsCan via the Data Liberation Initiative at academic institutions.

https://www150.statcan.gc.ca/n1/en/catalogue/98-401-X2016044.

## ACKNOWLEDGEMENTS

Reported COVID-19 cases were obtained from the Public Health Ontario Integrated Public Health Information System (iPHIS) via the Ontario COVID-19 Modelling Consensus Table and with approval from the University of Toronto Health Sciences Research Ethics Board (protocol no. 39253). A subset of iPHIS data (laboratory-confirmed cases only) were made available by the Ontario Ministry of Health and Long-Term Care (MOHLTC) to the Ontario Modelling Consensus Table on a daily basis. The analyses, conclusions, opinions and statements expressed herein are solely those of the authors and do not reflect those of the funding or data sources; no endorsement is intended or should be inferred.

SM is supported by a Tier 2 Canada Research Chair in Mathematical Modeling and Program Science. MM-G is supported by a Tier 2 Canada Research Chair in Population Health Modeling. SS is supported by a Tier 1 Canada Research Chair in Knowledge Translation and Quality of Care.

## CONFLICTS OF INTEREST TO DECLARE

None

## REFERENCES

1. Subedi R, Greenberg L, Turcotte M. COVID-19 mortality rates in Canada’s ethno-cultural neighbourhoods: c2020 [updated Oct 28, 2020; cited Jan 14, 2021]. Available from: https://www150.statcan.gc.ca/n1/pub/45-28-0001/2020001/article/00079-eng.htm.

2. Millet GA, Jones AT, Benkeser D, Baral SD, Mercer L, Beyrer C, et al. Assessing differential impacts of COVID-19 on black communities. Ann Epidemiol. 2020;47:37–44. doi:10.1016/j.annepidem.2020.05.003. PubMed Central PMCID: PMC32419766.

3. Sohn A, Phanuphak N, Baral S, Kamarulzaman A. Know your epidemic, know your response: understanding and responding to the heterogeneity of the COVID-19 epidemics across Southeast Asia. J Int AIDS Soc. 2020;23(7):e25557. doi:10.1002/jia2.25557.

4. Phaswana-Mafuya N, Shisana O, Gray G, Zungu N, Bekker L-G, Kuonza L, et al. The utility of 2009 H1N1 pandemic data in understanding the transmission potential and estimating the burden of COVID-19 in South Africa to guide mitigation strategies. S Afr Med J. 2020;110(7):576–7. doi:10.7196/SAMJ.2020.v110i7.14935.

5. Gonzalez-Reiche AS, Hernandez MM, Sullivan MJ, Ciferri B, Alshammary H, Obla A, et al. Introductions and early spread of SARS-CoV-2 in the New York City area. Science. 2020;369(65011):297–301. doi:10.1126/science.abc1917.

6. Wang L, Ma H, Yiu KCY, Calzavara A, Landsman D, Luong L, et al. Heterogeneity in testing, diagnosis and outcome in SARS-CoV-2 infection across outbreak settings in the Greater Toronto Area, Canada: an observational study. CMAJ Open. 2020;8(4):E627–E36. doi:10.9778/cmajo.20200213. PubMed Central PMCID: PMC33037070.

7. Ontario Ministry of Finance. Ontario Population Projections Update, 2019–2046: c2020 [cited Mar 24, 2021]. Available from: https://www.fin.gov.on.ca/en/economy/demographics/projections/.

8. Sundaram ME, Calzavara A, Mishra S, Kustra R, Chan AK, Hamilton MA, et al. The individual and social determinants of COVID-19 in Ontario, Canada: a population-wide study. medRxiv. 2020. doi:10.1101/2020.11.09.20223792.

9. Lee WC. Characterizing exposure-disease association in human populations using the Lorenz curve and Gini index. Stat Med. 1997;16(7):729–39. doi:0.1002/(sici)1097-0258(19970415)16:7<729::aid-sim491>3.0.co;2-a. PubMed Central PMCID: PMC9131761.

10. Elliott LJ, Blanchard JF, Beaudoin CM, Green CG, Nowicki DL, Matusko P, et al. Geographical variations in the epidemiology of bacterial sexually transmitted infections in Manitoba, Canada. Sex Transm Infect. 2002;78(Suppl 1):i139–44. doi:10.1136/sti.78.suppl_1.i139. PubMed Central PMCID: PMC12083433.

11. Kerani RP, Handcock MS, Handsfield HH, Holmes KK. Comparative geographic concentrations of 4 sexually transmitted infections. Am J Public Health. 2005;95(2):324–30. doi:10.2105/AJPH.2003.029413. PubMed Central PMCID: PMC15671471.

12. Mody A, Pfeifauf K, Geng EH. Using Lorenz curves to measure racial inequities. JAMA Netw Open. 2021;4(1):e2032696. doi:10.1001/jamanetworkopen.2020.32696.

13. Gomes M, Corder R, King J, Langwig K, Souto-Maior C, Carneiro J, et al. Individual variation in susceptibility or exposure to SARS-CoV-2 lowers the herd immunity threshold. medRxiv. 2020. doi:10.1101/2020.04.27.20081893.

14. Rodriguez J, Patón M,Acuña J. Prioritisation of population groups with the most interactions for COVID-19 vaccination can substantially reduce total fatalities. medRxiv. 2020. doi:10.1101/2020.10.12.20211094.

15. Chen Y, Glymour M, Riley A, Balmes J, Duchowny K, Harrison R, et al. Excess mortality associated with the COVID-19 pandemic among Californians 18–65 years of age, by occupational sector and occupation: March through October 2020. medRxiv. 2021. doi:10.1101/2021.01.21.21250266.

16. Bayati M, Feyzabadi VY, Rashidian A. Geographical disparities in the health of Iranian women: health outcomes, behaviors, and health-care access indicators. Int J Prev Med. 2017;8:11. doi:10.4103/ijpvm.IJPVM_67_16. PubMed Central PMCID: PMC28348721.

17. Farmer J, McLeod L, Siddiqi A, Ravghi V, Quiñoneza C. Examining changes in income-related oral health inequality in Canada: a population-level perspective. Can J Dent Hyg. 2016;50(2):65–71.

18. Neumayer E, Plümper T. Inequalities of income and inequalities of longevity: a cross-country study. Am J Public Health. 2016;106(1):160–5. doi:10.2105/AJPH.2015.302849. PubMed Central PMCID: PMC26562120.

19. Rashid MM, Roy TK, Singh BP. Socioeconomic health disparities in Bangladesh through Lorenz curve and Gini ratio: evidence from BDHS surveys. Int J Stat Anal. 2019;9(1):11–23. doi:10.37622/IJSA/9.1.2019.11-23

20. Martens PJ, Brownell M, Au W, MacWilliam L, Prior H, Schultz J, et al. Health inequities in Manitoba: is the socioeconomic gap widening or narrowing over time? Winnipeg: Manitoba Centre for Health Policy, 2010.

21. Equator Network. The Strengthening the Reporting of Observational Studies in Epidemiology (STROBE) Statement: guidelines for reporting observational studies: c2019 [cited Mar 31, 2021]. Available from: https://www.equator-network.org/reporting-guidelines/strobe/.

22. Statistics Canada. Toronto, C [Census subdivision], Ontario and Canada [Country] (table). Census Profile.: c2017 [updated Nov 29, 2017; cited Jan 14, 2021]. Available from: https://www12.statcan.gc.ca/census-recensement/2016/dp-pd/prof/index.cfm?Lang=E.

23. Statement by the Minister of Health on the first presumptive confirmed travel-related case of new coronavirus in Canada [Internet]. 2020; Jan 25, 2020 [cited Jan 14, 2021]. Available from:.https://www.canada.ca/en/public-health/news/2020/01/statement-by-the-minister-of-health-on-the-first-presumptive-confirmed-traveled-related-case-of-new-coronavirus-in-canada.html

24. Public Health Ontario. COVID-19 in Ontario: January 15, 2020 to January 12, 2021 2021.

25. Statistics Canada. Dissemination area: detailed definition: 2018 [updated Sep 17, 2018; cited Jan 14, 2021]. Available from: https://www150.statcan.gc.ca/n1/pub/92-195-x/2011001/geo/da-ad/def-eng.htm.

26. Statistics Canada. Postal CodeOM Conversion File (PCCF). 2019.

27. 2016 Census of Population. Census Profile - Age, Sex, Type of Dwelling, Families, Households, Marital Status, Language, Income, Immigration and Ethnocultural Diversity, Housing, Aboriginal Peoples, Education, Labour, Journey to Work, Mobility and Migration, and Language of Work for Canada, Provinces and Territories, Census Divisions, Census Subdivisions and Dissemination Areas (File: 98-401-X2016044) [Internet]. 2017. Available from: https://www150.statcan.gc.ca/n1/en/catalogue/98-401-X2016044.

28. Statistics Canada. Census Family Status and Household Living Arrangements (13), Household Type of Person (9), Age (12) and Sex (3) for the Population in Private Households of Canada, Provinces and Territories, Census Metropolitan Areas and Census Agglomerations, 2016 and 2011 Census. 2019.

29. Martin CA, Jenkins DR, Minhas JS, Gray LJ, Tang J, Williams C, et al. Socio-demographic heterogeneity in the prevalence of COVID-19 during lockdown is associated with ethnicity and household size: Results from an observational cohort study. EClinicalMedicine. 2020;25:100466. doi:10.1016/j.eclinm.2020.100466.

30. van den Broek-Altenburg Em, Atherly AJ, Diehl SA, Gleason KM, Hart VC, MacLean CD, et al. Jobs, housing, and mask wearing: cross-sectional study of risk factors for COVID-19. City of Toronto. COVID-19JMIR Public Health Surveill. 2021;7(1):e24320. doi:10.2196/24320.

31. Karaye IM, Horney JA. The impact of social vulnerability on COVID-19 in the U.S.: an analysis of spatially varying relationships. Am J Prev Med. 2020;59(3):317–25. doi:10.1016/j.amepre.2020.06.006.

32. Elliott S, Leon S. Crowded housing and COVID-19: impacts and solutions: c2020.[updated Jul 24, 2020; cited Jan 21, 2021]. Available from: https://www.wellesleyinstitute.com/healthy-communities/crowded-housing-and-covid-19-impacts-and-solutions/.

33. Statistics. National Occupational Classification (NOC) 2016 Version 1.3: c2016 [updated 2020 Jan 13; cited Mar 18, 2021]. Available from: https://www23.statcan.gc.ca/imdb/p3VD.pl?Function=getVD&TVD=1267777.

34. BMJ. 11. Correlation and regression: c2021 [cited Mar 19, 2021]. Available from: https://www.bmj.com/about-bmj/resources-readers/publications/statistics-square-one/11-correlation-and-regression.

35. Government of Ontario. Reopening Ontario in stages: c2020 [cited Jan 6, 2021]. Available from: https://www.ontario.ca/page/reopening-ontario-stages.

36. Jefferies S, French N, Gilkison C, Graham G, Hope V, Marshall J, et al. COVID-19 in New Zealand and the impact of the national response: a descriptive epidemiological study. Lancet Public Health. 2020;5(11):E612–E23. doi:10.1016/S2468-2667(20)30225-5. PubMed Central PMCID: PMC33065023.

37. Otu A, Ahinkorah B, Ameyaw E, Seidu A, Yaya S. One country, two crises: what Covid-19 reveals about health inequalities among BAME communities in the United Kingdom and the sustainability of its health system? Int J Equity Health. 2020;19:189. doi:10.1186/s12939-020-01307-z.

38. Drefahl S, Wallace M, Mussino E, Aradhya S, Kolk M, Brandén M, et al. A population-based cohort study of socio-demographic risk factors for COVID-19 deaths in Sweden. Nat Commun. 2020;11:5097. doi:10.1038/s41467-020-18926-3.

39. Wang L, Moqueet N, Simkin A, Knight J, Ma H, Lachowsky N, et al. Mathematical modelling of the influence of serosorting on the population-level HIV transmission impact of pre-exposure prophylaxis. AIDS. 2021. doi:10.1097/QAD.0000000000002826.

40. Wellman B, Hogan B. Connected lives: the project. In: Networked Neighborhoods: The Connected Community in Context. London: Springer; 2006.

41. van Houtven CH, DePasquale N, Nb C. Essential long-term care workers commonly hold second jobs and double- or triple-duty caregiving roles. J Am Geriatr Soc. 2020;68(8):1657–60. doi:10.1111/jgs.16509. PubMed Central PMCID: PMC32338767.

42. Connor J, Madhavan S, Mokashi M, Amanuel H, Johnson NR, Pace LE, et al. Health risks and outcomes that disproportionately affect women during the Covid-19 pandemic: a review. Soc Sci Med. 2020;266:113364. doi:10.1016/j.socscimed.2020.113364.

43. Thakur N, Lovinsky-Desir S, Bime C, Wisnivesky JP, Celedón JC. The structural and social determinants of the racial/ethnic disparities in the U.S. COVID-19 pandemic: what’s our role? Am J Resp Crit Care Med. 2020;202(7):943–9. doi:10.1164/rccm.202005-1523PP

44. Brandén M, Aradhya S, Kolk M, Härkönen J, Drefahl S, Malmberg B, et al. Residential context and COVID-19 mortality among adults aged 70 years and older in Stockholm: a population-based, observational study using individual-level data. Lancet Healthy Longev. 2020;1(2):e80–e8. doi:10.1016/S2666-7568(20)30016-7.

45. Madewell Z, Yang Y, Longini I, Halloran E, Dean N. Household transmission of SARS-CoV-2: a systematic review and meta-analysis. JAMA Netw Open. 2020;3(12):e2031756. doi:10.1001/jamanetworkopen.2020.31756(

46. City of Toronto. COVID-19: status of cases in Toronto: c2020 [cited Jan 13, 2021]. Available from: https://www.toronto.ca/home/covid-19/covid-19-latest-city-of-toronto-news/covid-19-status-of-cases-in-toronto/.

47. Weill JA, Stigler M, Deschenes O, Springborn MR. Social distancing responses to COVID-19 emergency declarations strongly differentiated by income. Proc Natl Acad Sci USA. 2020;117(33):19658–60. doi:10.1073/pnas.2009412117. PubMed Central PMCID: PMC32727905.

48. Yang J, Allen K, Bailey A. What cellphone mobility data can teach us about why lockdown might not be working, and what to expect from the holidays. Toronto Star. 2020 Dec 13, 2020.

49. Kustra R. Regional (Public Health Unit) mobility trends by workplace. Google Open Data. [personal communications]. 2021.

50. Science Advisory and Modelling Consensus Tables. Update on COVID-19 projections [presentation]. 2021.

51. Wrigley-Field E, Kiang M, Riley A, Barbieri M, Chen Y-H, Duchowny K, et al. Geographically-targeted COVID-19 vaccination is more equitable than age-based thresholds alone. medRxiv. 2021. doi:10.1101/2021.03.25.21254272;

52. Allen U. Reducing the impact of COVID-19 on Black communities in Canada: building confidence and decreasing vaccine hesitancy. RSC COVID-19 Series. 2021 Mar 3, 2021:82.

53. Goodnough A, Hoffman J. The wealthy are getting more vaccinations, even in poorer neighborhoods. The New York Times. 2021 Feb 2, 2021.

54. City of Toronto Board of Health. Response to COVID-19 - March 2021 Update. Mar 22, 2021 ed2021.

55. Health Commons Solutions Lab. Community-driven testing strategies for COVID-19 [presentation]. 2021.

56. Belluz J. Social distancing is a luxury many can’t afford. Vermont actually did something about it. voxcom. 2020 Nov 19, 2020.

57. Cevik M, Baral S, Crozier A, Cassell J. Support for self-isolation is critical in covid-19 response. BMJ. 2021;372:224. doi:10.1136/bmj.n224.

58. Abbasi K. Why vaccinating staff and supporting self-isolating people are national emergencies. BMJ. 2021;372:239. doi:10.1136/bmj.n239.

59. Pichler S, Wen K, Ziebarth N. COVID-19 emergency sick leave has helped flatten the curve in the United States. Health Affairs. 2020;39(2). doi:10.1377/hlthaff.2020.00863.

60. Public Health Agency of Canada. From risk to resilience: an equity approach to COVID-19. 2020.

61. City of Toronto Board of Health. Response to COVID-19 - January 2021 Update. Jan 5, 2021 ed2021.

