## Appendices for "Increasing concentration of COVID-19 by socioeconomic determinants and geography in Toronto, Canada: an observational study"

**FIGURES**

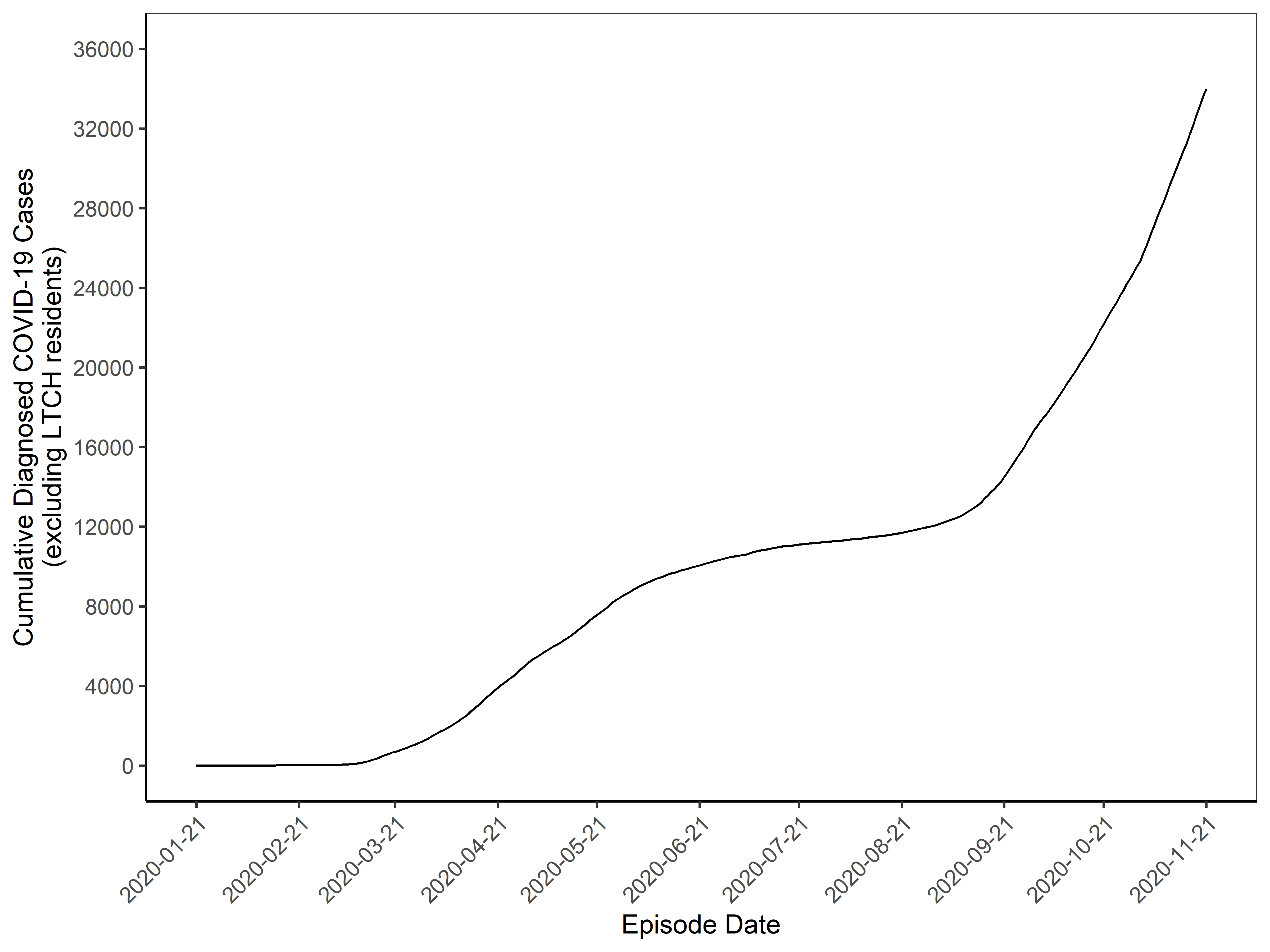

**Figure A1. Cumulative epidemic curve of diagnosed COVID-19 cases in Toronto, Canada (January 21, 2020 to November 21, 2020).** Total of 33,992 cases of COVID-19, excluding LTCH residents, were observed during this period.
LTCH: long-term care homes

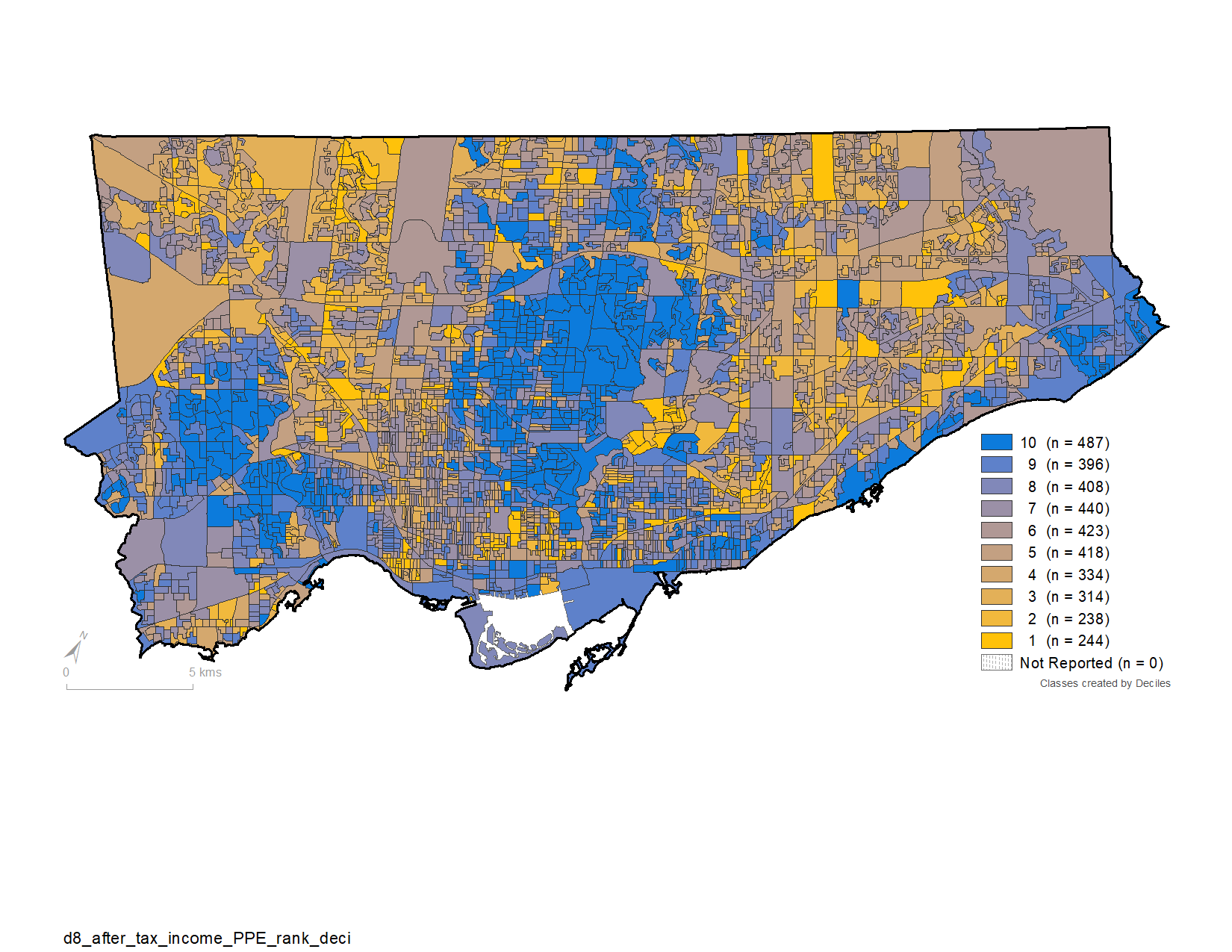

A) Household income

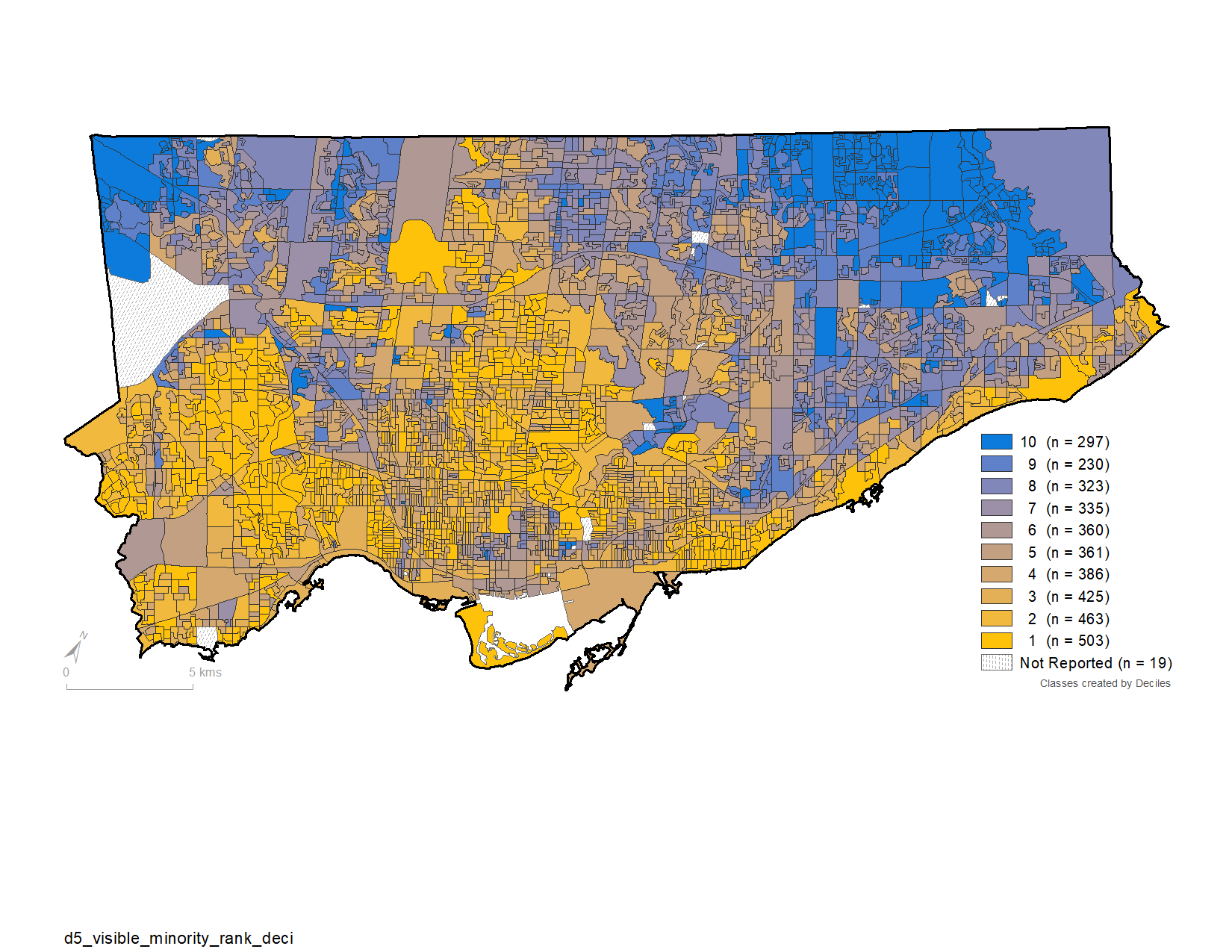

B) Visible minority

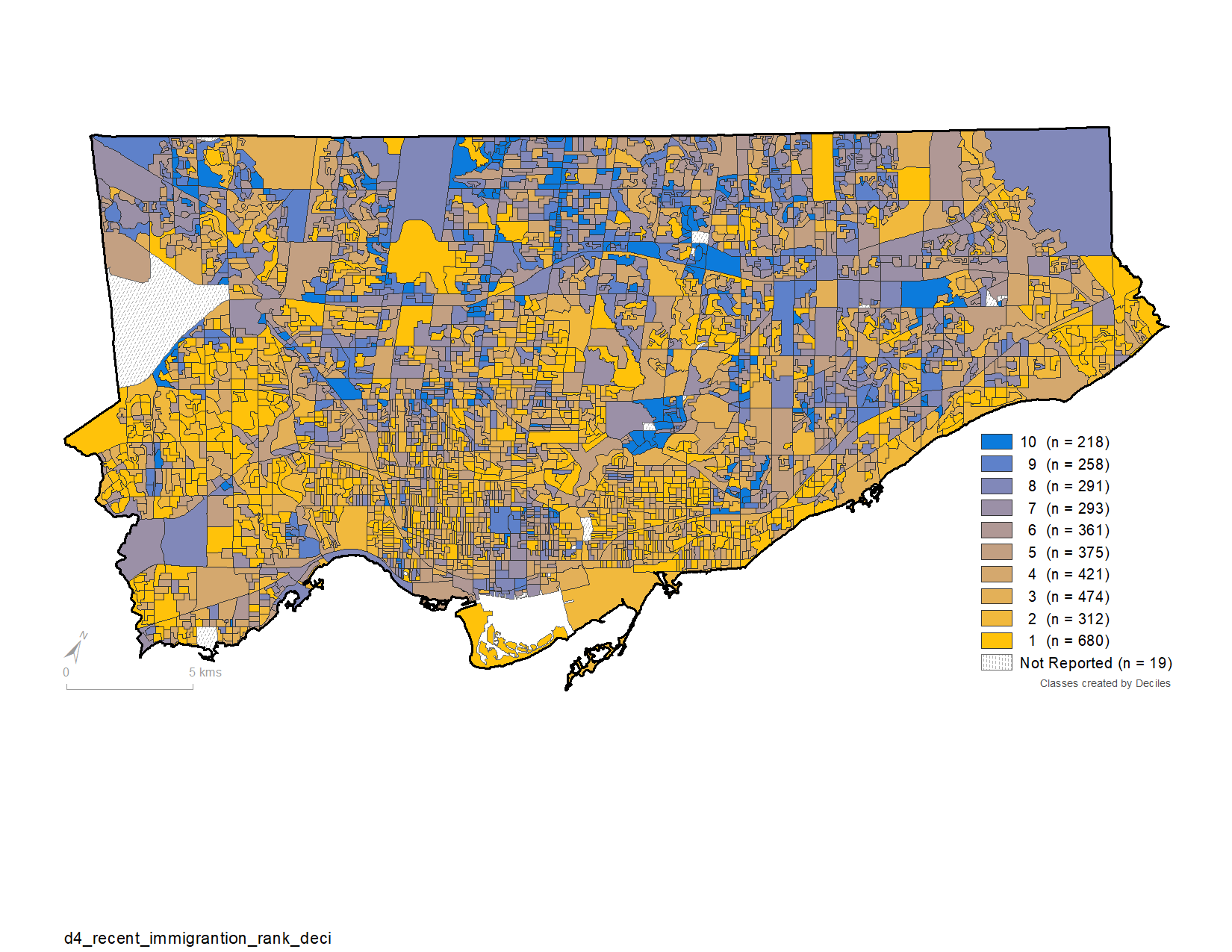

C) Recent immigration

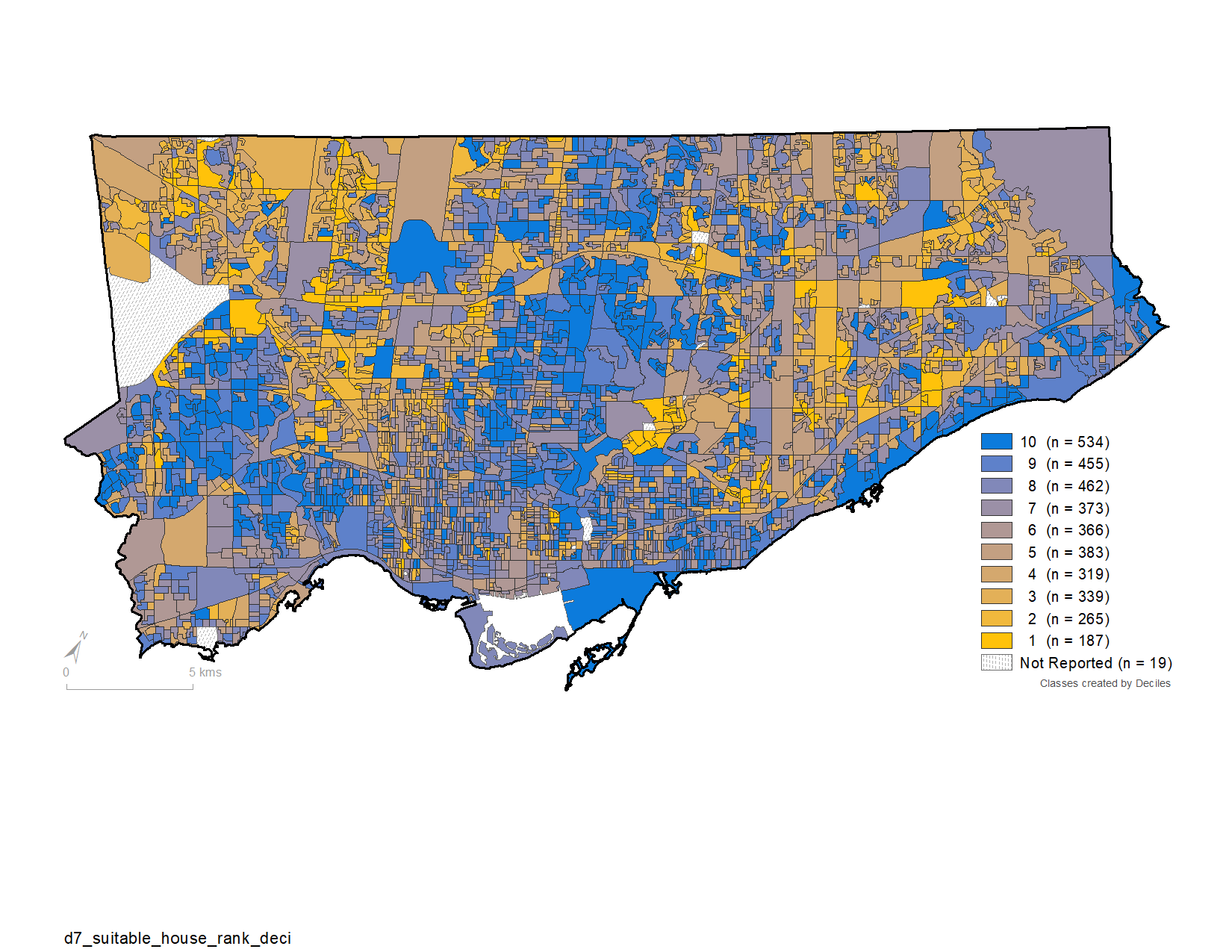

D) Suitable housing

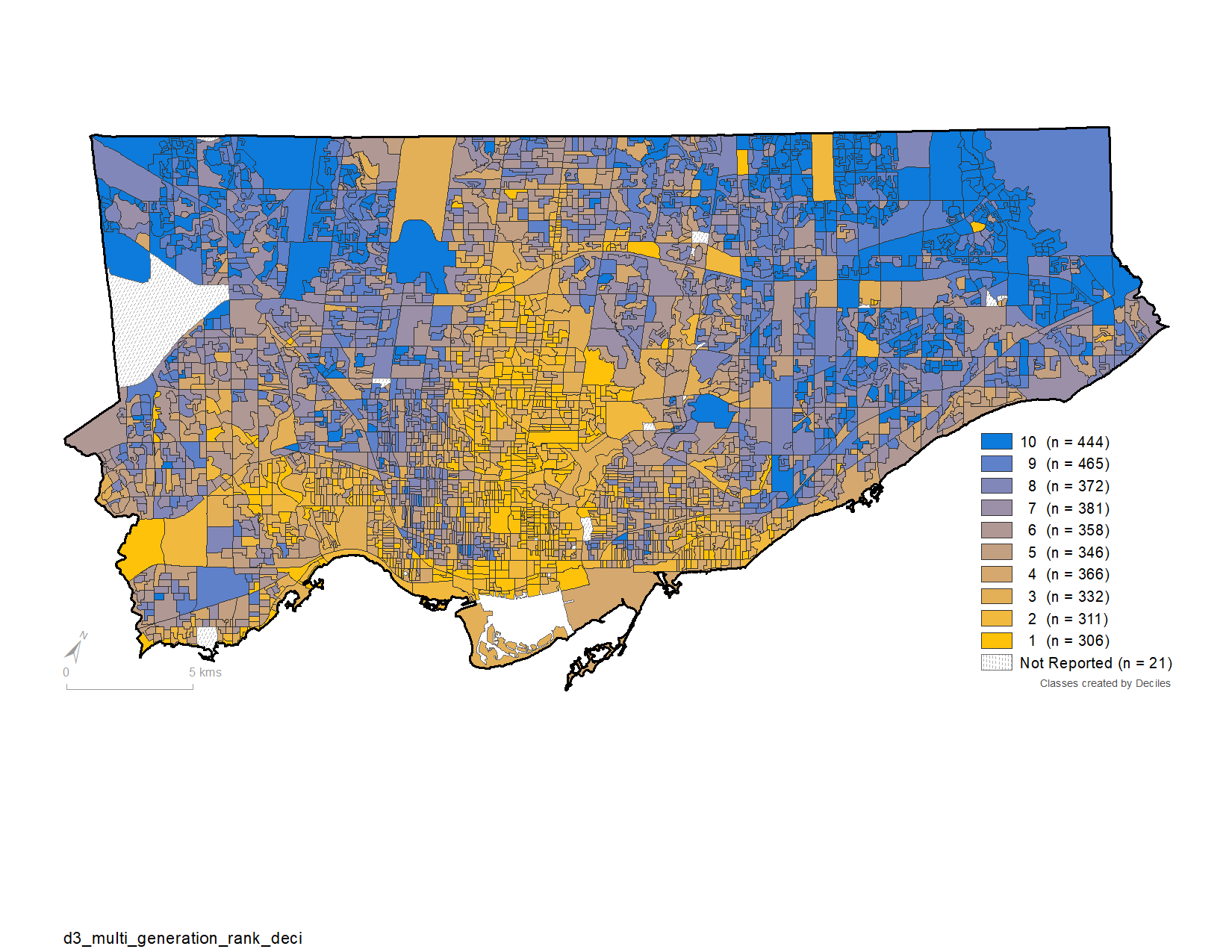

E) Multi-generational households

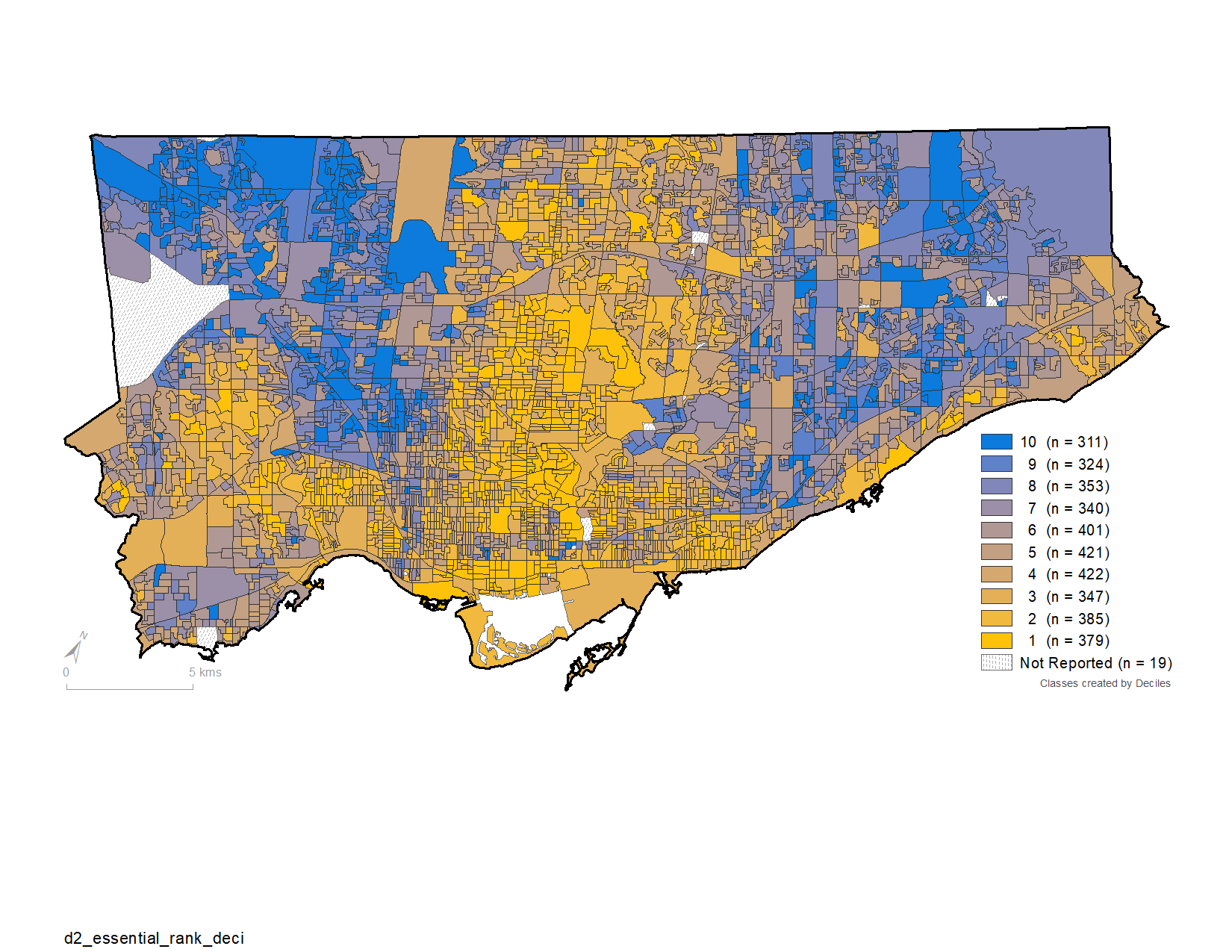

F) Essential workers

**Figure A2. Distribution of social determinant deciles by dissemination areas (DA) in the City of Toronto, Canada.** Heat maps of DAs within the City of Toronto public health unit depicting lowest to highest deciles with respect to: household income (A); % visible minority (B); % recent immigration (C); % suitable housing (D); % multi-generational households (E); and % essential workers (F). The lowest decile (decile 1) represents areas with the lowest income (A), lowest proportion of visible minority (B), lowest proportion of recent immigrants (C), the lowest proportion of homes deemed suitable housing (D), lowest proportion of multi-generational households (E), and fewest essential workers (F). The highest decile (decile 10) represents areas with the highest income (A), highest proportion of visible minority (B), highest proportion of recent immigrants (C), highest proportion of homes deemed suitable housing (D), highest proportion of multi-generational households (E), and highest prevalence of essential workers (F).

DA: dissemination area

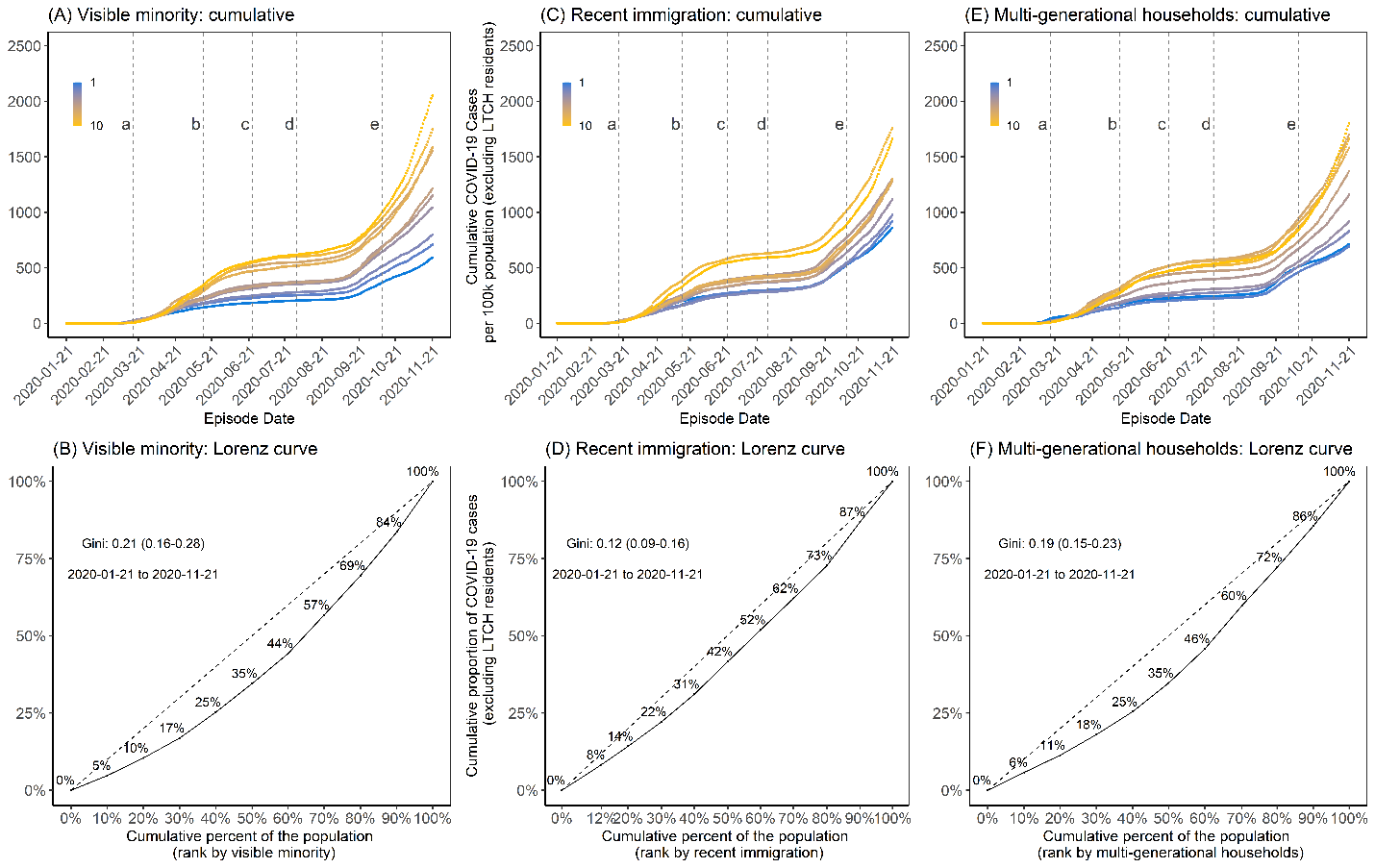

**Figure A3. Cumulative epidemic curves, and cumulative Lorenz curve by three social determinants (visible minority, recent immigration, and multi-generational households).**  Panels A and B represent visible minority deciles where the lowest decile (decile 1) represents the lowest proportion of visible minority and decile 10 represents the highest proportion of visible minority. Panels D and E represent recent immigration deciles where the lowest decile (decile 1) represents lowest proportion of recent immigrants; and the highest decile (decile 10) represents highest proportion of recent immigrants. Panel G and H represent multi-generational households where the lowest decile represents DAs with the lowest proportion of multi-generational households and the highest decile (decile 10) represents the highest proportion of multi-generational households. The Lorenz curves (Panel B, D, and F) depict the concentration of cases by each determinant. The time-periods are: (a) March 17, 2020, start of shutdown; (b) May 14, 2020, start of stage 1 re-opening; (c) June 24, 2020, start of stage 2 re-opening; (d) July 31, 2020, start of stage 3 re-opening; and (e) October 10, 2020, start of modified stage 2.
DA: dissemination area; LTCH: long-term care homes

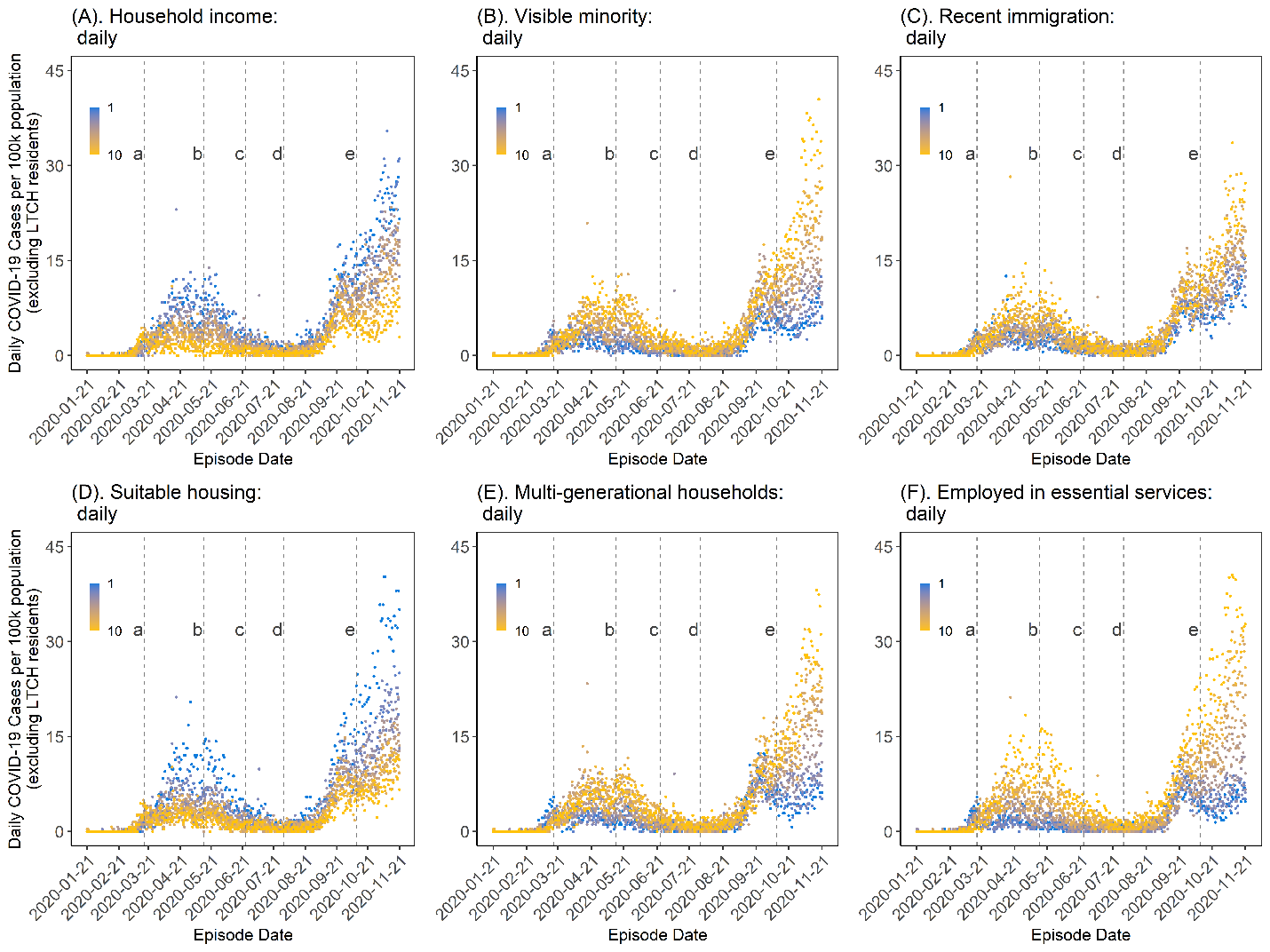

**Figure A4. Daily epidemic curves by six social determinants.** Income values described are per-person equivalent and after-tax (A). The time-periods are: (a) March 17, 2020, start of shutdown; (b) May 14, 2020, start of stage 1 re-opening; (c) June 24, 2020, start of stage 2 re-opening; (d) July 31, 2020, start of stage 3 re-opening; and (e) October 10, 2020, start of modified stage 2 re-opening.
DA: dissemination area; LTCH: long-term care homes

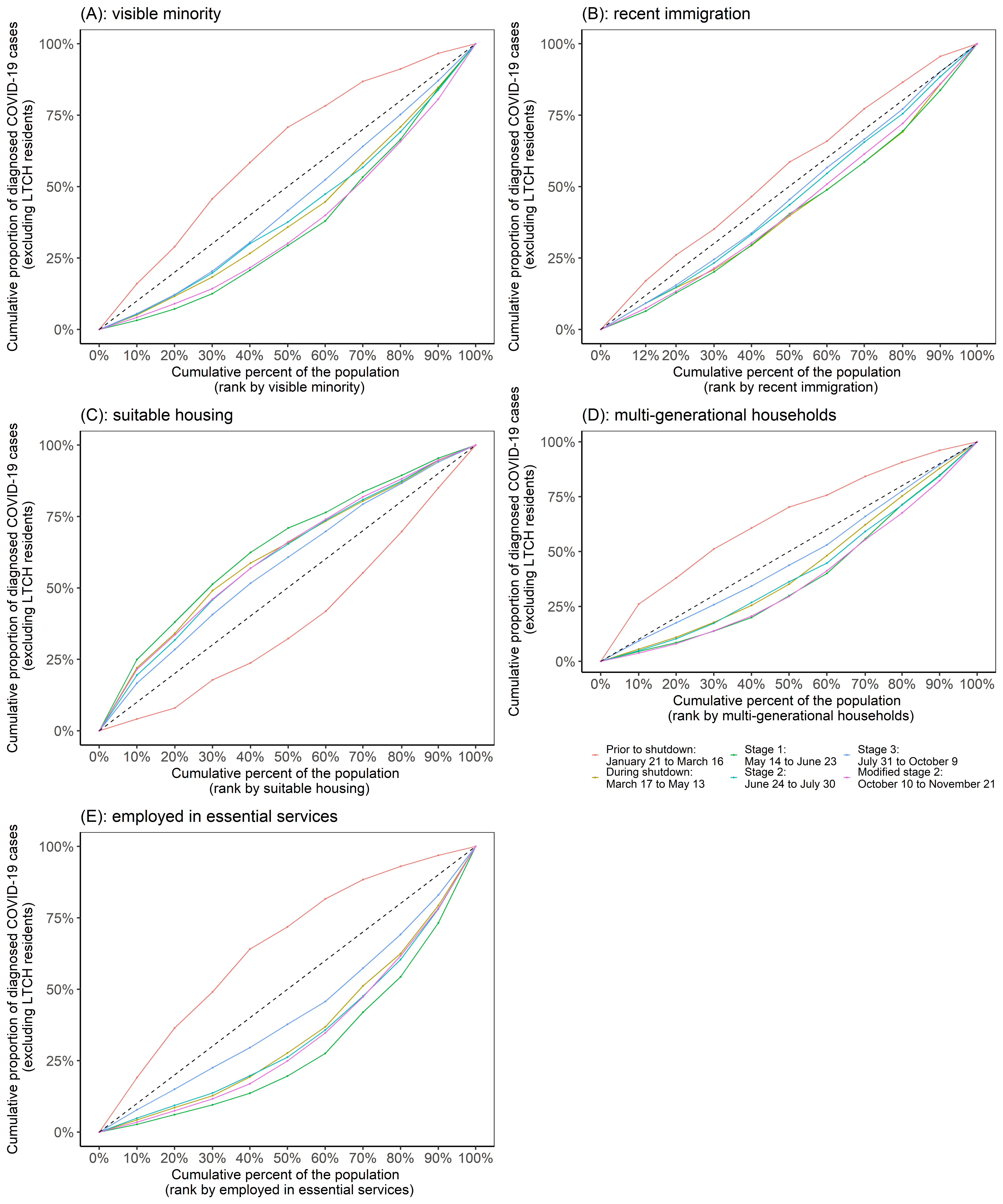

**Figure A5. Lorenz curve and Gini coefficient of COVID-19 cases over time by five social determinants (visible minority, recent immigration, suitable housing, multi-generational households, and essential workers) in Toronto, Canada (January 21, 2020 to November 21, 2020).** The dashed line is the line of equality. The coloured solid lines represent the time periods associated with the stages of intervention: prior to shutdown (January 21, 2020 to March 16, 2020); during shutdown (March 17, 2020 to May 13, 2020); stage 1 re-opening (May 14, 2020 to June 23, 2020); stage 2 re-opening (June 24, 2020 to July 30, 2020); stage 3 re-opening (July 31, 2020 to October 9, 2020); and modified stage 2 re-opening (October 10, 2020 to November 21, 2020).
LTCH: long-term care homes

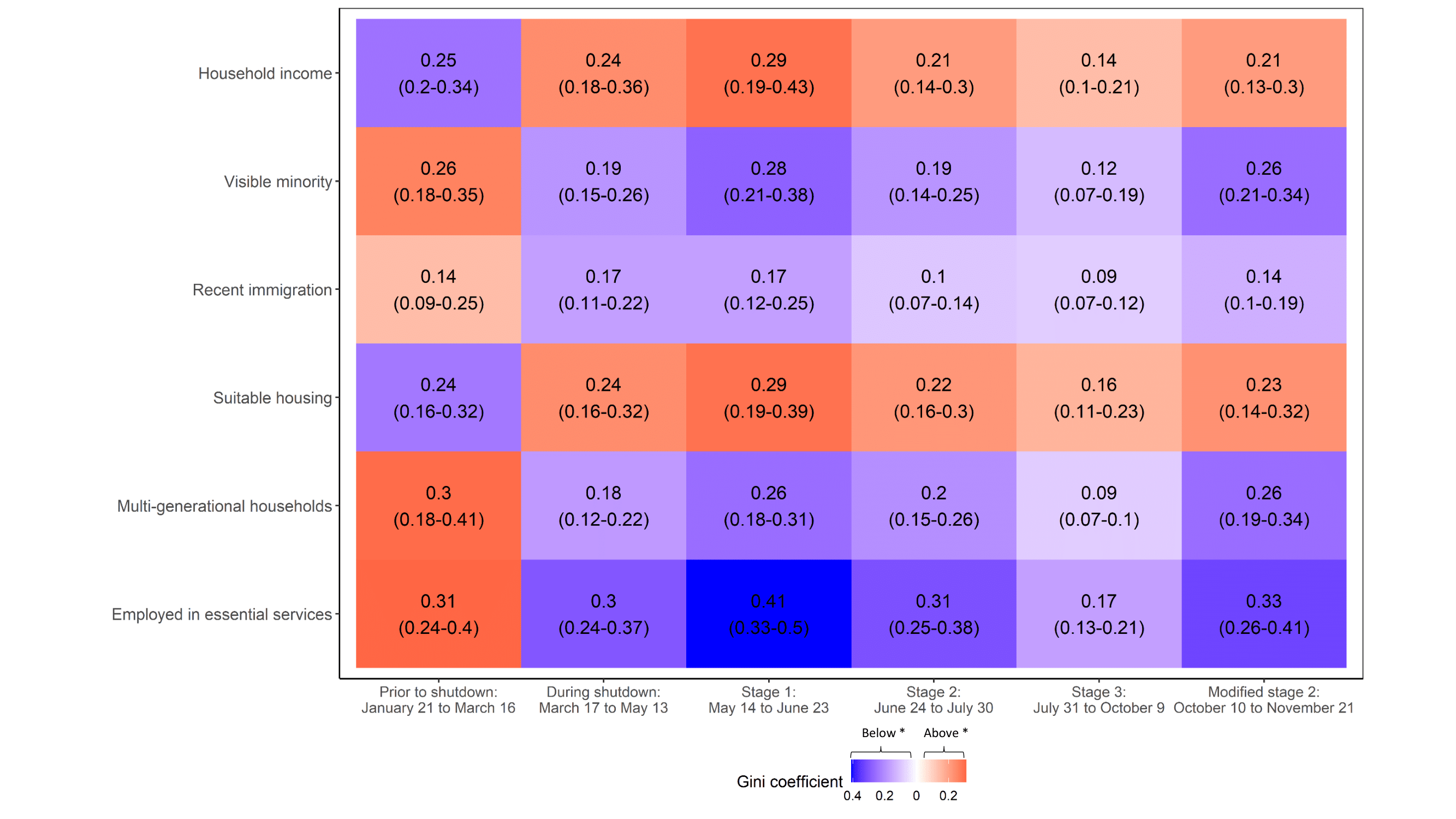

**Figure A6. Gini coefficient of COVID-19 cases over time by social determinants in Toronto, Canada (January 21, 2020 - November 21, 2020).** A heat map depicting the Gini coefficient of COVID-19 cases by household income, visible minority, recent immigration, suitable housing, multi-generational households, and essential worker. Gini coefficient above the line of equality is depicted in red and below the line of equality is depicted in purple. The time-periods are: (a) March 17, 2020, start of shutdown; (b) May 14, 2020, start of stage 1 re-opening; (c) June 24, 2020, start of stage 2 re-opening; (d) July 31, 2020, start of stage 3 re-opening; and (e) October 10, 2020, start of modified stage 2 re-opening.

**TABLES**

**Table A1. Social determinants of health – Variables from Statistics Canada 2016 Census of Population**

| Measure (*Source*)*^*^* | Definition of indicator | Notes^†^ (27, 63) |
| --- | --- | --- |
| Population size  (100% *of census sample*) | Total population count of a Dissemination Area | In this measure and where required, Dissemination Area population counts are adjusted (reduced) to remove residents of Long Term Care Homes (LTCH)^‡^. |
| **Socio-demographic** |  |  |
| Household income  (*100% of census sample*)^§^ | Decile rank of a Dissemination Area’s average total after-tax income, weighted by population | After-tax income is calculated for each household from the income for all household members. Calendar year 2015 is the reference period for all income variables in the 2016 Census. Single-person equivalent is used to account for households of different sizes. To limit variations in the cost of living, the ranking is calculated exclusively from DAs within the City of Toronto. |
| % recent immigration  (*25% of census sample*) | Numerator: Number of persons who immigrated to Canada in the 5 year period between 2011 and 2016  Denominator: Total population within the Dissemination Area | 2016 Census Dictionary states: 'Immigrant' refers to a person who is, or who has ever been, a landed immigrant or permanent resident. Such a person has been granted the right to live in Canada permanently by immigration authorities.  2016 Census Dictionary states: 'Period of immigration' refers to the period in which the immigrant first obtained landed immigrant or permanent resident status. |
| % visible minority  (*25% of census sample*) | Numerator: Number of persons who belong to visible minority groups  Denominator: Total population within the Dissemination Area | Visible minority groups are defined by the Employment Equity Act: "persons, other than Aboriginal peoples, who are non-Caucasian in race or non-white in colour". 2016 Census Dictionary states: “The visible minority population consists mainly of the following groups: South Asian, Chinese, Black, Filipino, Latin American, Arab, Southeast Asian, West Asian, Korean and Japanese.” |
| **Dwelling-related** |  |  |
| % suitable housing  (*25% of census sample*) | Numerator: Number of private households^\|\|^ living in dwellings that have “enough bedrooms for the size and composition of the household.” [2016 Census Dictionary]  Denominator: Total number of private households within the Dissemination Area | The National Occupancy Standard (NOS) is used to classify the suitability of accommodations. A suitable household is defined as "households where the required number of bedrooms based on the National Occupancy Standard (NOS) does not exceed the reported number of bedrooms in the dwelling.” The number of required bedrooms is determined using the following criteria:  1. A maximum of two persons per bedroom.  2. Household members, of any age, living as part of a married or common-law couple share a bedroom with their spouse or common-law partner.  3. Lone-parents, of any age, have a separate bedroom.  4. Household members aged 18 or over have a separate bedroom - except those living as part of a married or common-law couple.  5. Household members under 18 years old of the same sex share a bedroom - except lone-parents and those living as part of a married or common-law couple.  6. Household members under 5 years old of the opposite sex share a bedroom if doing so would reduce the number of required bedrooms. This situation would arise only in households with an odd number of males under 18, an odd number of females under 18, and at least one female and one male under the age of 5.  <https://www23.statcan.gc.ca/imdb/pUtil.pl?Function=getNote&Id=141809&NT=01> |
| multi-generational households  (*100% of census sample*) | Numerator: Number of persons who live in households where “at least one person [in the household is] living with a child and a grandchild.” [2016 Census Dictionary]  Denominator: Total of all persons who are classified by family status and household living arrangements | This measure is a count of persons whose households are described by a specific living arrangement. All persons in the same household are counted separately. In the numerator for this measure, family status includes persons who are married spouses, common-law partners, lone parent families, and the child(ren) of these persons. Persons living alone, with other relatives, or with non-relatives only are additionally included in the denominator count of persons. Couples can be opposite or same sex. |
| **Occupation-related** |  |  |
| % essential services not amenable to remote working  (25% *of census sample*) | Numerator: Number of persons in the labor force who have occupations in one of the following categories: Manufacturing/utilities, Trades/transport/equipment operators, Sales/services, Health, Resources/agriculture/production  Denominator: Total labor force population aged 15 years and over in private households in the Dissemination Area | Occupations are assigned according to the National Occupancy Classification (2016). Occupation was chosen over “Industry” to better represent the type of work performed and skill-level required by a population rather than the industry that provides the employment. Numerators may be defined separately (“or”) or added together in different combination sets (“and”). “Labor Force” is all persons in private households aged 15 years and older who were either employed or unemployed during the week of Sunday, May 1 to Saturday, May 7, 2016. |

* “Sample” refers to the short-form Census questionnaire (*100%* sample) or to the long-form questionnaire, received by a random sample of households (*25%* sample). It is mandatory for recipients to respond to the questionnaires. Statistical inferences for the entire population are drawn from the subset of responses of the long-form questionnaire; these inferences are reported in the tabulated values provided by Statistics Canada. Note that income information was collected solely from administrative data sources (*100%* sample) and were not part of either questionnaire.

^†^Additional details about variable definitions may be included the Census Dictionary; please refer to Statistics Canada’s Dictionary for the 2016 Census of Population for complete definitions. Some definitions provided here are taken verbatim from source.

^‡^ Due to reporting methods used by CCM+, case counts among “Long-Term Care Residents” may also include cases that are reported for residents of “nursing home[s] or other chronic care facility[ies]”. Adjustments in population counts described here only include adjustments to Dissemination Areas that have one (or more) LTCH facility identified by the Ontario Ministry of Health. The adjustments are made by subtracting the total number of beds in the facility from the population count of the DA.

^§^ Income deciles for the City of Toronto / Toronto Public Health Unit were tabulated by ICES from data contained in PCCF+ (version 7B) and adjusted for population size. **Ref: Statistics Canada. 2018. Postal Code Conversion File Plus (PCCF+) Version 7B, Reference Guide. November 2018 Postal codes**.

^||^ Where referenced, “household” refers to a “private household”. The 2016 Census Dictionary states: ”Private household” refers to a person or group of persons who occupy the same dwelling and do not have a usual place of residence elsewhere in Canada or abroad.”

**Table A2. Shutdown and reopening stages in Ontario**

| **Shutdown and stages of reopening^[1]^** | **Public and social gatherings^[2]^** | **Industry services and retail** | **Outdoor, recreational and seasonal activities** | **Care, community, and household services** | **Academic and teaching institutions** |
| --- | --- | --- | --- | --- | --- |
| Shutdown^[3]^ (64, 65) | General guidelines   - Limit for outdoor organized public events and social gatherings (excluding those within same household): 5 people - Every person shall stay-at-home except for essential reasons - Physical distancing of at least two metres away from others outside of their direct household   Permitted with requirements   - Funeral service with limit of 10 people^[4]^ (66) | Open for essential businesses including   - Supply chains - Retail and wholesale of food, pet food and supplies, and household consumer products - Liquor stores - Gas stations and fuel providers - Motor repair - Hardware stores - Pharmaceutical services - Office supplies and services - Safety supply - Restaurants and food facilities^[5]^ - Hotels and shared rental units - Support and maintenance services - Telecommunications and IT infrastructure/providers - Transportation providers and supporting services - Manufacturers - Agriculture and food production - Construction - Financial activities - Resources - Environmental services - Utilities and community services - Communications industries - Research - Justice sector - Other businesses^[6]^ - Business regulators and inspectors | All non-essential businesses closed; no essential businesses in this category | Open for essential businesses including   - Health care and seniors care and social services - Child care centres^[7]^ | Publicly-funded elementary and secondary closed starting March 14, 2020 (67, 68)  Suspension or modification of classes at post-secondary institutions starting mid-March 2020^[8]^ (69) |
| **Reopening**^[9]^ | | | | | |
| Stage 1 of reopening^[10]^ (70, 71) | General guidelines   - Social gathering limit: 5 people - Physical distancing of least two metres away from others outside of their direct household - Distancing of vehicle by at least 2 metres - Limit outings and public gatherings as per emergency orders | Reopen/resumption with restrictions and limitations, subject to conditions   - Construction - Retail located outside of shopping malls^[11]^ - Fitting rooms at retail settings - Motor vehicle dealerships - Media operations - Non-essential professional services - Emissions inspection facilities | Reopen/resumption with restrictions and limitations, subject to conditions   - Golf driving ranges - Recreational services at marinas - Rod and gun clubs - Cycling tracks - Sport activity for individual/single competitors^[12]^ - Ontario Parks with limited backcountry camping (72) | Reopen/resumption with restrictions and limitations, subject to conditions   - Health care services - Supporting services for surgeries and procedures - In-person counselling - Veterinary services - Libraries - Domestic services^[13]^ - Maintenance, repair and property management services | Closed (73)   - Public and private schools - Licensed child care centres - EarlyON programs |
| Stage 2 of reopening^[14],^^[15]^ (72) | General guidelines   - Social gathering limit: 10 people - Physical distancing of at least two metres away from others outside of their direct household - Non-essential in-person gatherings of any size continue to be strongly discouraged   Permitted with requirements   - Small outdoor events - Reopening of places of worship ^[16]^ - Venues not otherwise restricted can open to conduct wedding ceremonies, funerals and similar gatherings | Reopen/resumption with restrictions and limitations, subject to conditions   - Shopping centres, malls and markets - Photography studios and services - Film and television production activities - Indoor and outdoor tour and guide services - Restaurants, bars, food trucks and other food and drink establishments for dining in outdoor areas | Reopen/resumption with restrictions and limitations, subject to conditions   - Indoor and outdoor swimming pools and outdoor splash pads and wading pools ^[17]^ - Outdoor-only recreational facilities that operate low-contact attractions and activities - Indoor recreational activities^[18]^ - Ontario Parks campgrounds and private campgrounds for recreational vehicle, car camping and all other types of camping - Outdoor team sports ^[19]^ - Drive-in and drive-through venues - Attractions and heritage institutions^[20]^ | Reopen/resumption with restrictions and limitations, subject to conditions   - Personal care services^[21]^ - Libraries ^[22]^ - Community centres^[23]^ | Reopen/resumption with restrictions and limitations, subject to conditions   - Post-secondary education^[24]^ (74) |
| Stage 3 of reopening^[25]^ (75) | General guidelines   - Gathering limits^[26]^: 50 people for indoor^[27]^ and 100 people for outdoor - Physical distancing of at least two metres with people from outside their households or social circles   Permitted with requirements   - Indoors gathering for religious services, rites or ceremonies, and wedding ceremonies or funeral services | Reopen/resumption with restrictions and limitations, subject to conditions   - Indoor dine-in at restaurants, bars, concession stands, and other food and drink establishments^[28],^^[29]^ | Reopen/resumption with restrictions and limitations, subject to conditions   - Exhibits with high-contact surfaces - On-site services at libraries may - Concerts, artistic events, theatrical productions, performances, and movie theatres ^[30]^ - Indoor recreational activities ^[31]^ - Casinos and charitable gaming establishments^[32]^ - Drive-in and drive-through venues^[33]^ - Facilities for sports and recreational fitness activities^[34]^ - Outdoor playgrounds and play structures - Team sports and live sporting events^[35]^ | Reopen/resumption with restrictions and limitations, subject to conditions   - Libraries^[36]^ - Personal care services^[37]^ | Elementary and secondary schools permitted to reopen for 2020-21 school year starting September 8, 2020 (76) |
| Modified stage 2 of reopening^[38]^ (62, 77) | General guidelines^[39]^   - Social gatherings and organized public events with limit of 10 people indoors, 25 people outdoors - Tour and guide services with limit of 10 people indoors, 25 people outdoors - Open houses with limit of 10 people indoors   General guidelines updated^[40]^   - Meeting and event spaces with limit of 10 people indoors, 25 people outdoors^[41]^ - Wedding receptions with limit of 10 people indoors, 25 people outdoors - Places of worship remain open | Closed^[42]^   - Indoor food and drink service in restaurants and other food and drink establishments - Real estate open houses^[43]^   Closed with exceptions^[44]^   - Conference and convention centres^[45]^ | Closed^[46]^   - Indoor gyms and fitness centres - Casinos and other gaming establishments - Cinemas - Performing arts centres and venues - Spectator areas in racing venues - Interactive exhibits or exhibits with high risk of personal contact   Closed with exceptions   - Team sports^[47]^ | Remain open   - Child care centres   Closed   - Personal care services where face coverings must be removed for service | Remain open   - Schools^[48]^   Restrictions   - In-person teaching and instruction^[49]^ |

**Table A3. Characteristics of the study population and dissemination area, City of Toronto, Canada**

|  |  | Total number of dissemination areas (N=3,702) | | Total study population  (N = 2,731,571) |
| --- | --- | --- | --- | --- |
| Study population | Population size of dissemination area, (median, IQR)^*^ | N of dissemination areas | % of dissemination areas | % of population |
| Overall | 450 (446 - 768) | 3,702 | 100.0 | 100.0 |
| 1st quintile | 416 (365 - 459) | 1,376 | 37.2 | 20.1 |
| 2nd quintile | 522 (523 - 587) | 991 | 26.8 | 20.2 |
| 3rd quintile | 739 (676 - 842) | 710 | 19.2 | 19.8 |
| 4th quintile | 1,223 (1,069 – 1,434) | 429 | 11.6 | 19.9 |
| 5th quintile | 2,430 (2,042 – 3,035) | 196 | 5.3 | 20.0 |
| Dissemination areas with zero cases^†^ | N/A | 404 | 10.9 | 8.1 |

^*^Based on Census Canada estimates (27)

^†^Excluding cases diagnosed among LTCH residents; laboratory-confirmed cases only from January 21, 2020 to November 21, 2020.
LTCH: long-term care homes; IQR: interquartile range.

**Table A4. Mean and median values and distribution of dissemination areas of social determinants by deciles**

| **Social Determinant** | | **Value** | | **Dissemination Areas (DA)** | | |
| --- | --- | --- | --- | --- | --- | --- |
| **Variable** | **Decile** | **Mean** | **Median (IQR)** | **N** | **% of DAs** | **Cumulative per-capita cases** |
| Household income | 1 | 22724.3 | 24123.0 (19663.2 - 26164.5) | 244 | 6.6 | 1942.2 |
|  | 2 | 30499.5 | 30731.0 (29401.0 – 31615.0) | 238 | 6.4 | 1938.1 |
|  | 3 | 34637.0 | 34626.0 (33644.5 – 35716.0) | 314 | 8.5 | 1435.9 |
|  | 4 | 38058.4 | 38098 (37435.5 - 38684.2) | 334 | 9.0 | 1387.2 |
|  | 5 | 41261.5 | 41192 (40406.8 - 42129.8) | 418 | 11.3 | 1282.9 |
|  | 6 | 44979.6 | 44981 (44057.5 – 45872.0) | 423 | 11.4 | 1217.6 |
|  | 7 | 48800.9 | 48793.0 (47731.0 – 49853.0) | 440 | 11.9 | 1079.1 |
|  | 8 | 53734.2 | 53390.5 (52122.5 - 55466.5) | 408 | 11.0 | 916.6 |
|  | 9 | 61562.8 | 61406.5 (59179.5 – 63672.0) | 396 | 10.7 | 686.6 |
|  | 10 | 87749.8 | 79183.0 (71615.0 – 95666.0) | 487 | 13.2 | 556.0 |
| % visible minority | 1 | 11.3 | 11.8 (8.8 - 14.2) | 503 | 13.6 | 592.2 |
|  | 2 | 19.9 | 19.9 (18.1 - 21.9) | 463 | 12.5 | 709.2 |
|  | 3 | 27.0 | 26.9 (25.2 - 28.9) | 425 | 11.5 | 798.4 |
|  | 4 | 35.2 | 35.2 (32.9 - 37.2) | 386 | 10.4 | 1043.3 |
|  | 5 | 45.5 | 45.6 (42.9 – 48.0) | 361 | 9.8 | 1149.7 |
|  | 6 | 55.8 | 55.8 (53.1 - 58.3) | 360 | 9.7 | 1213.6 |
|  | 7 | 65.6 | 65.5 (62.8 - 68.2) | 335 | 9.0 | 1552.2 |
|  | 8 | 75.5 | 75.3 (73.0 - 77.7) | 323 | 8.7 | 1583.3 |
|  | 9 | 84.1 | 84.0 (82.3 - 85.7) | 230 | 6.2 | 1746.9 |
|  | 10 | 93.9 | 93.9 (91.0 - 96.8) | 297 | 8.0 | 2052.2 |
|  | Missing | Missing | Missing | 19 | 0.5 | 5340.7 |
| % recent immigration | 1 | 0 | 0 (0 - 0) | 680 | 18.4 | 860.9 |
|  | 2 | 1.7 | 1.7 (1.5 - 1.9) | 312 | 8.4 | 919.8 |
|  | 3 | 2.4 | 2.4 (2.2 - 2.6) | 474 | 12.8 | 975.5 |
|  | 4 | 3.5 | 3.5 (3.2 - 3.7) | 421 | 11.4 | 1116.8 |
|  | 5 | 4.6 | 4.6 (4.3 - 4.9) | 375 | 10.1 | 1303.0 |
|  | 6 | 6.1 | 6.1 (5.7 - 6.5) | 361 | 9.8 | 1285.2 |
|  | 7 | 7.8 | 7.8 (7.3 - 8.2) | 293 | 7.9 | 1278.1 |
|  | 8 | 9.8 | 9.7 (9.1 - 10.4) | 291 | 7.9 | 1298.6 |
|  | 9 | 13.1 | 12.9 (12.0 - 14.2) | 258 | 7.0 | 1758.2 |
|  | 10 | 21.0 | 19.6 (17.2 - 22.4) | 218 | 5.9 | 1661.8 |
|  | Missing | Missing | Missing | 19 | 0.5 | 5340.7 |
| % suitable housing | 1 | 62.2 | 64.2 (58.2 - 67.7) | 187 | 5.1 | 2543.8 |
|  | 2 | 76.0 | 76.6 (73.7 - 78.2) | 265 | 7.2 | 1486.2 |
|  | 3 | 82.4 | 82.6 (81.2 - 83.8) | 339 | 9.2 | 1622.9 |
|  | 4 | 86.3 | 86.3 (85.4 - 87.1) | 319 | 8.6 | 1297.4 |
|  | 5 | 89.4 | 89.5 (88.9 – 90.0) | 383 | 10.3 | 1075.6 |
|  | 6 | 91.6 | 91.7 (91.1 - 92.1) | 366 | 9.9 | 982.9 |
|  | 7 | 93.6 | 93.6 (93.1 – 94.0) | 373 | 10.1 | 1022.1 |
|  | 8 | 95.4 | 95.3 (94.9 - 95.9) | 462 | 12.5 | 807.7 |
|  | 9 | 97.1 | 97.1 (96.8 - 97.4) | 455 | 12.3 | 902.5 |
|  | 10 | 100.2 | 100 (100 - 100) | 534 | 14.4 | 687.4 |
|  | Missing | Missing | Missing | 19 | 0.5 | 5340.7 |
| % multi-generational households | 1 | 0.5 | 0.5 (0 - 0.9) | 306 | 8.3 | 713.4 |
|  | 2 | 1.7 | 1.7 (1.4 – 2.0) | 311 | 8.4 | 693.9 |
|  | 3 | 2.8 | 2.8 (2.5 - 3.1) | 332 | 9.0 | 830.4 |
|  | 4 | 4.3 | 4.3 (3.9 - 4.8) | 366 | 9.9 | 919.4 |
|  | 5 | 5.9 | 5.9 (5.5 - 6.2) | 346 | 9.3 | 1158.7 |
|  | 6 | 7.6 | 7.6 (7.2 – 8.0) | 358 | 9.7 | 1372.2 |
|  | 7 | 9.4 | 9.4 (8.9 - 9.9) | 381 | 10.3 | 1700.5 |
|  | 8 | 11.6 | 11.6 (11.0 - 12.2) | 372 | 10.0 | 1581.2 |
|  | 9 | 15.1 | 14.9 (13.9 - 16.3) | 465 | 12.6 | 1662.3 |
|  | 10 | 23.5 | 22.3 (19.7 – 26.0) | 444 | 12.0 | 1802.5 |
|  | Missing | Missing | Missing | 21 | 0.6 | 3068.6 |
| % essential workers | 1 | 20.2 | 20.9 (18.7 - 22.9) | 379 | 10.2 | 600.8 |
|  | 2 | 27.1 | 27.1 (25.7 - 28.4) | 385 | 10.4 | 617.1 |
|  | 3 | 31.6 | 31.6 (30.6 - 32.5) | 347 | 9.4 | 618.0 |
|  | 4 | 36.6 | 36.6 (35.3 - 37.9) | 422 | 11.4 | 744.0 |
|  | 5 | 42.4 | 42.4 (40.9 - 43.9) | 421 | 11.4 | 981.6 |
|  | 6 | 48.3 | 48.1 (46.8 – 50.0) | 401 | 10.8 | 1121.6 |
|  | 7 | 53.8 | 53.9 (52.6 - 55.1) | 340 | 9.2 | 1592.1 |
|  | 8 | 58.6 | 58.5 (57.3 - 59.9) | 353 | 9.5 | 1598.8 |
|  | 9 | 64.1 | 64 (62.7 - 65.6) | 324 | 8.8 | 1990.8 |
|  | 10 | 72.4 | 71.4 (69.2 - 74.4) | 311 | 8.4 | 2580.4 |
|  | Missing | Missing | Missing | 19 | 0.5 | 5340.7 |

DA: dissemination area; IQR: interquartile range

**References (continuation from main text)**

27.	2016 Census of Population. Census Profile - Age, Sex, Type of Dwelling, Families, Households, Marital Status, Language, Income, Immigration and Ethnocultural Diversity, Housing, Aboriginal Peoples, Education, Labour, Journey to Work, Mobility and Migration, and Language of Work for Canada, Provinces and Territories, Census Divisions, Census Subdivisions and Dissemination Areas (File: 98-401-X2016044) [Internet]. 2017. Available from: <https://www150.statcan.gc.ca/n1/en/catalogue/98-401-X2016044>.

62.	Government of Ontario. Archived - Reopening Ontario in stages: c2020 [updated Nov 3, 2020; cited Mar 12, 2020]. Available from: https://www.ontario.ca/page/reopening-ontario-stages.

63.	Statistics Canada. 2016 Census Dictionary. 2017.

64.	Government of Ontario. Ontario extends emergency declaration to stop the spread of COVID-19: c2020 [cited Mar 12, 2021]. Available from: https://news.ontario.ca/en/release/56523/ontario-extends-emergency-declaration-to-stop-the-spread-of-covid-19.

65.	Government of Ontario. Emergency Management and Civil Protection Act - ONTARIO REGULATION 82/20. March 24, 2020 to April 2, 2020 ed2020.

66.	Government of Ontario. Ontario Regulation REG2020.0205.e. 2020.

67.	Government of Ontario OotP. Statement from Premier Ford, Minister Elliott, and Minister Lecce on the 2019 Novel Coronavirus (COVID-19). Mar 12, 2020 ed2020.

68.	Government of Ontario. School closures extended to keep students, staff and families safe: c2020 [updated Apr 26, 2020; cited Mar 17, 2021]. Available from: https://news.ontario.ca/en/release/56776/school-closures-extended-to-keep-students-staff-and-families-safe.

69.	Freeman J. GTA universities, colleges suspending classes amid pandemic. CTV News. 2020 2020 Mar 13.

70.	Government of Ontario. Archived - A framework for reopening our province: Stage 1: c2020 [cited Mar 12, 2021]. Available from: https://www.ontario.ca/page/framework-reopening-our-province-stage-1#section-2.

71.	Government of Ontario. Ontario extends emergency orders to keep people safe: c2020 [updated May 19, 2020; cited Mar 16, 2021]. Available from: https://news.ontario.ca/en/release/56968/ontario-extends-emergency-orders-to-keep-people-safe.

72.	Government of Ontario. Archived - A framework for reopening our province: Stage 2: c2020 [cited Mar 12, 2021]. Available from: https://www.ontario.ca/page/framework-reopening-our-province-stage-2.

73.	Health and safety top priority as schools remain closed [Internet]. 2020; 2020 May 19 [cited Mar 17, 2021]. Available from: https://news.ontario.ca/en/release/56971/health-and-safety-top-priority-as-schools-remain-closed

74.	Ontario unveils a plan to reopen postsecondary education [Internet]. 2020; 2020 Jun 10 [cited Mar 17, 2021]. Available from: https://news.ontario.ca/en/release/57150/ontario-unveils-a-plan-to-reopen-postsecondary-education

75.	Government of Ontario. Archived - A framework for reopening our province: Stage 3: c2020 [updated Nov 3, 2020; cited Mar 12, 2021]. Available from: https://www.ontario.ca/page/framework-reopening-our-province-stage-3#:~:text=In%20Stage%203%2C%20more%20restrictions,necessary%20to%20keep%20everyone%20safe.

76.	Government of Ontario. Archived - Approach to reopening schools for the 2020-2021 school year: c2020 [cited Mar 17, 2021]. Available from: https://www.ontario.ca/page/approach-reopening-schools-2020-2021-school-year.

77.	Government of Ontario. Ontario moving additional region to modified stage 2. Oct 16, 2020 ed2020.

1. Each stage outlines the newly added list of activities allowed to resume in addition to the businesses and activities reopened in the previous stage.
2. Gathering limits and general public health and safety guidelines apply to all businesses, events, and activities. Exceptions are detailed in the footnotes.
3. Closure of non-essential businesses – with the exception of essential businesses mentioned, all other businesses are closed.
4. Gathering limit of 5 people is exempted for funerals.
5. Only for delivery or takeaway.
6. Other businesses include rental and leasing services, businesses providing mailing and delivery services, laundry service providers, professional services, businesses providing funeral and related goods and services, land registration/real estate agent services and moving services, security services, staffing services, support of safe operations of residences and essential businesses, businesses that provide for health and welfare of animals, child care services for essential workers, businesses providing cheque cashing services.
7. Permitted for home child care services of up to six children and child care centres for essential workers.
8. Decisions were made independently by each school.
9. Establishments permitted to reopen must follow proper health and safety protocols (i.e., physical distancing, proper cleaning and disinfection). Businesses, services and locations not mentioned in current or previous stages of reopening remain closed.
10. Toronto Public Health started stage 1 on May 19, 2020.
11. Must set restrictions to allow physical distancing (e.g., limiting number of customers in store, booking appointments, encouraging curbside pickup or deliveries).
12. Includes indoor and outdoor non-team sport competitions that can be played while maintaining physical distancing and without spectators.
13. Indoor and outdoor household services that can adhere to public health guidelines.
14. Toronto Public Health started stage 2 on June 24, 2020.
15. Businesses can open to offer other permitted services even if its primary services is a restricted activity. For example, facial salons are permitted to open for other services.
16. Attendance is limited to 30% of building capacity.
17. Must have no access to high-contact aquatic features. Waterparks, wave pools, and water slides are not open.
18. Only indoor driving ranges and rod and gun clubs.
19. For training only and with no scrimmages or games and with limited access to facilities (i.e., no locker rooms or change rooms, etc.).
20. Interactive and high-contact exhibits, amusement parks, water parks and conference centres remain closed.
21. Restrictions include: a) prohibit of services that tend to a customer’s face; b) continued closure of steam rooms, saunas, bath houses; c) closure of baths, hot tubs, floating pools, and sensory deprivation pods with exceptions for therapeutic purposes.
22. Limited on-site services (e.g., computer access and contactless book pickup and drop-off).
23. Limited or modified on-site programs and services (e.g., physically distanced programs and services for in-person counselling, computer access, tutoring, etc.).
24. In-person instruction permitted for students in essential, frontline, and high labour market demand areas (e.g., nursing, personal support workers, etc.). Publicly assisted colleges and universities, Indigenous Institutes, private career colleges and other postsecondary education institutions may participate in this voluntary reopening.
25. Toronto Public Health started stage 3 on July 31, 2020.
26. People at their place of work do not count towards gathering limits.
27. Indoor gathering limits apply to events that are fully or partially indoors. Indoor gathering size cannot be increased by combining with outdoor event.
28. As of September 26, 2020, all food and drink establishments must: 1) stop selling alcohol at 11:00pm; 2) prohibit consumption of alcohol between 12:00am and 9:00am; 3) close by 12:00am; 4) remain closed until 5:00am (with exceptions of takeout and delivery)
29. Buffet-style service must remain closed.
30. Impermeable barrier (e.g., plexiglass) is required between audience and singers and players of brass or wind instruments.
31. Karaoke is permitted only outside of private karaoke rooms.
32. Table games must remain closed.
33. Not subject to gathering limits.
34. Physical distancing must be maintained with exception of playing in team sport or if needed for personal training purposes. Gathering limits apply to areas containing weights or exercise machines (indoor gathering limit) and in classes or organized activities (indoor and outdoor gathering limits). Gathering limits do not apply in all other areas (e.g., pools, tennis courts and rinks). Steam rooms and saunas must remain closed.
35. Team sports that require prolonged or deliberate body contact between players are not yet permitted, unless it could be modified to prevent prolonged or deliberate physical contact. Leagues cannot contain more than 50 participants.
36. For all on-site services given library materials are disinfected or quarantined before recirculation.
37. All services that tend to a customer’s face are permitted. Oxygen bars, bath houses, steam rooms and saunas must remain closed.
38. Modified stage 2 includes additional public health measures to those implemented in stage 2.
39. As of October 10, 2020.
40. As of October 13, 2020
41. Exemptions include for government operations and delivery of government services.
42. As of October 10, 2020.
43. In-person showing by appointments only.
44. As of October 13, 2020.
45. With exception of where there are courts.
46. As of October 10, 2020.
47. Except for training sessions. No games or scrimmages permitted.
48. Before-school and after-school programs remain open.
49. Capacity limit mentioned under general guidelines for social gatherings and organized public events apply, with certain exemptions, including for schools, universities, colleges, etc.
